# Recent antipsychotics use associated with elevated risk of Parkinson’s disease

**DOI:** 10.64898/2026.06.19.26356070

**Authors:** Lee Neilson, Ryan Carnahan, Susan Duffy, Vicki Kijewski, Nandakumar Narayanan, Jacob Simmering

## Abstract

**Introduction:** Parkinson’s disease is a neurodegenerative disease affecting motor and cognitive function that has a major impact on society. Epidemiological evidence has suggested that the incidence of PD may be increasing; however, the underlying etiology is unclear. Here, we investigated the role of increasingly used antipsychotics in the diagnosis of PD.

**Methods:** We harnessed Merative insurance claims databases to conduct a case-control study of 65,275 new cases of PD and 652,364 age-, sex-, and time-matched controls. We estimated associations between exposure and duration of use for antipsychotics adjusted for important confounders using fixed effects logistic regression. We performed sensitivity analyses stratified by the level of D2 receptor inhibition to assess dose-response relationships; a lagged exposure analysis to address confounding by indication; analysis assessing exposure of other psychiatric medications without significant D2 inhibition (bupropion, trazodone, and Z-drugs); analysis assessing exposure of non-psychiatric medications with significant (metoclopramide) or no D2 inhibition (ondansetron).

**Results:** We found cases with PD had elevated odds of antipsychotic exposure. Longer durations of exposure and greater affinity for the D2 receptor were associated with greater associations with PD. There was a dose-response relationship between D2 inhibition activity and increased odds of PD for a similar duration of exposure. There was a dose response relationship between duration of metoclopramide and the odds of PD; however, there was no such relationship between the non-D2 inhibiting control medications. The association between exposure to an antipsychotic and increased odds of PD was present even when the first exposure was 10 years prior to the PD diagnosis date.

**Conclusion:** If these results are causal, antipsychotic use may explain up to 2.4% of all cases of PD. Given the increasing rate of use of these medications, and the concurrent increasing age-adjusted incidence of PD, there is an urgent need for further investigation into this association and greater awareness of the potential risks of these medications in older adults.

## INTRODUCTION

Parkinson’s disease (PD) incidence has been increasing for several decades at a rate faster than can be explained by changing demographics alone (1–4). While there is emerging evidence of environmental exposures in the development of PD (5–7), other contributors may include the use of medications that are known to block dopamine receptors, chiefly antipsychotics (8). It is well known that D2 blockade may cause extrapyramidal symptoms (9). However, recent evidence suggests that these dopamine-blocking medications may be associated with elevated risk of degenerative PD (10,11) and increased morbidity in people with PD (12). These common and widely prescribed medications (13,14) causing long-term D2 blockade are an intriguing possible contributor to the increasing incidence of PD. We tested the hypothesis in a case-control study using a large database of insurance claims.

## METHODS

### Data Source

Our data source was the Merative Marketscan Research Databases Commercial Claims and Encounters (CCAE) and Medicare Coordination of Benefits (MDCR) databases for the years 2001-2024. These databases contain claims with detailed diagnosis, procedure, and pharmaceutical information for over 220 million unique enrollees in private health insurance plans, including employer-sponsored, retiree, and Medicare supplemental plans in the United States. Since our exposure of interest is pharmaceutical use, we required that the enrollee had active prescription drug coverage during the lookback period.

### Case Definition

We defined a case of PD as a person of any age having 1) at least two dates with a claim with a diagnosis of PD in any setting (ICD-9-CM 332.0, ICD-10-CM: G20.*), 2) the first and last dates with a PD diagnoses were at least 90 days apart, and 3) the first and last observed PD events (diagnosis or dispensing claim for levodopa) occurred at least 365 days apart. We further refined to incident cases of PD by requiring at least 365 days of enrollment prior to the first observed PD event. We excluded any patient with a secondary parkinsonism diagnosis (ICD-9-CM 332.1, ICD-10-CM: G21.*) occurring on or after the first observed PD event date to reduce the potential inclusion of drug-induced parkinsonism in our cases.

### Control Identification

For each case, we identified up to 10 enrollees who were never diagnosed with PD, treated with levodopa, or diagnosed with secondary parkinsonism and who were born in the same year, of the same sex, and enrolled for the same period of time as the PD case. We required the enrollment period to match to account for potential changes in diagnostic standards or other medical practice over time. Fewer than 10 matches would occur in rare instances where there were fewer than 10 eligible control enrollees. Cases with zero eligible controls were excluded from the analysis.

### Exposures

The time period of interest, the lookback period, is the time from the start of enrollment in Marketscan until the day prior to the PD diagnosis date. Antipsychotic dispensing events during this lookback period served as our exposures of interest. We identified all active ingredients in the WHO Collaborating Center for Drug Statistics Methodology Anatomical Therapeutic Chemical (ATC) (15) class N05A (Antipsychotics) using the National Library of Medicine’s RxMix tool (16). We identified National Drug Code numbers for all medications currently or ever licensed in the United States in the RED Book (17) with at least one of the active ingredients identified through RxMix. All medications are described in **Table 1**.

**Table 1:** Antipsychotics Medications Used in Study. Tier is the system of stratification we developed for the D2 Ki with Tier 1 medications having a D2 Ki <= 1 nM, Tier 2 1 nM < Ki < 10 nM, Tier 3 10 nM <= Ki < 100, and Tier 4 having Ki >= 100. We prefer this classification over generation as it better describes the D2 affinity of the compound. Observed dispensing events are the number of times this medication was included on a dispensing claim in the sample.

| Medication | Generation | D2 Receptor Ki (nM) | Tier | Observed Dispensing Events | Observed Days Supplied | Median Days Supplied (IQR) |
| --- | --- | --- | --- | --- | --- | --- |
| Chlorpromazine | First | 2.00 | 2 | 1,274 | 194,193 | 10 (5 – 30) |
| Droperidol | First | 0.25 | 1 | 8 |  |  |
| Fluphenazine | First | 0.54 | 1 | 200 | 104,527 | 180 (60 – 516) |
| Haloperidol | First | 2.00 | 2 | 2,113 | 491,658 | 56 (24 – 185) |
| Loxapine | First | 10.00 | 3 | 72 | 33,075 | 179.5 (30 – 510) |
| Mesoridazine | First | 19.00 | 3 | 10 | 3,558 | 105 (67.5 – 573) |
| Molindone | First | 63.00 | 3 | 13 | 8,947 | 414 (58 – 1,020) |
| Perphenazine | First | 1.40 | 2 | 853 | 659,400 | 420 (90 - 1,110) |
| Pimozide | First | 0.65 | 1 | 58 | 19,721 | 120 (50 – 487.5) |
| Prochlorperazine | First | 4.00 | 2 | 17,866 | 425,832 | 8 (5 – 15) |
| Thioridazine | First | 10.00 | 3 | 260 | 197,626 | 390 (59.25 – 1,087.5) |
| Thiothixene | First | 1.40 | 2 | 172 | 152,388 | 495 (77 – 1,230) |
| Trifluoperazine | First | 1.30 | 2 | 213 | 195,777 | 450 (90 – 1,500) |
| Aripiprazole | Second | 0.95 | 1 | 4,417 | 1,789,601 | 210 (60 – 540) |
| Asenapine | Second | 1.30 | 2 | 117 | 35,040 | 90 (30 – 307) |
| Clozapine | Second | 431.00 | 4 | 95 | 91,151 | 417 (59.5 – |
|  |  |  |  |  |  | 60.5) |
| Iloperidone | Second | 3.30 | 2 | 23 | 5,437 | 90 (30 – 165) |
| Lurasidone | Second | 0.99 | 1 | 239 | 72,603 | 150 (30 – 366.5) |
| Olanzapine | Second | 72.00 | 3 | 6,598 | 3,027,768 | 196 (60 – 570) |
| Paliperidone | Second | 4.00 | 2 | 174 | 60,229 | 180 (31.75 – 448.25) |
| Quetiapine | Second | 567.00 | 4 | 10,655 | 4,760,174 | 180 (45 – 570) |
| Risperidone | Second | 4.90 | 2 | 8,359 | 3,293,674 | 180 (60 – 480) |
| Ziprasidone | Second | 4.00 | 2 | 1,027 | 435,555 | 150 (30 – 502) |
| Brexpiprazole | Third | 0.30 | 1 | 123 | 34,844 | 120 (30 – 390) |
| Cariprazine | Third | 1.60 | 2 | 78 | 20,335 | 120 (5 – 30) |
| Lumateperone | Third | 32.00 | 3 | 2 | 90 | 45 (37.5 – 52.5) |
| Pimavanserin | Other | 300.00 | 4 | 7 | 1,500 | 90 (45 – 360) |

We defined two measures of exposure: did the enrollee ever have a pharmacy claim for one or more of the medications (ever use) and, in aggregate over all observed dispensing events, how many days of medications were supplied across these claims (duration).

### Primary Analysis

We sought to reduce this confounding unaddressed by the case-control matching using fixed effects logistic regression. We include a fixed effect for each case/control strata to account for matching process used to select cases and controls and, by extension, the different durations of lookback, temporal trends in medical care, sex, and age. We included covariates to adjust for health care utilization rates (rates of inpatient encounters and outpatient encounters during the lookback period), health complexity (average number of unique diagnoses per day with an outpatient claim), general health and comorbidities (the 30 Elixhauser/AHRQ comorbidity flags), and specific adjustment for psychiatric conditions (Clinical Classification Software (CCS) codes 651-664). We cluster our standard errors by the case/control strata to account for the matching of cases and controls.

We estimated a model for each of our three primary outcomes associating PD diagnosis with ever use and days supplied of antipsychotics. Ever use was binary (never user versus ever user) while days supplied were categorized into the levels “never supplied,” and 9 levels of duration of exposure among the exposed.

### Sensitivity Analyses

We defined seven sensitivity analyses to better describe the potential for a causal relationship between antipsychotic exposure and the odds of PD.

First, we demonstrate a dose-response pattern exploiting the fact that different antipsychotics have different affinity for the D2 receptor. We divide the antipsychotic medications into four tiers based on the D2 receptor Ki (tier 1 <= 1 nM, tier 2 > 1 nM and < 10 nM, tier 3 >= 10 nM and < 100 nM, tier 4 >= 100 nM) as reported in the PDSP database (18,19). The D2 Ki values used in our analysis are included in **Table 1**. We estimated the adjusted association with ever use and duration of exposure for each of the four tiers of Ki affinity.

Second, we address confounding due to possible prodromal symptoms triggering antipsychotic use or pharmacological unmasking of subclinical PD through a time-since-first-dispensing analysis. We calculated the time between the first observed dispensing for an antipsychotic and the PD index date. We categorized this interval into 17 bins, with bin width selected to contain approximately 500 PD cases. We estimated the association between these exposure interval bins and a diagnosis of PD while controlling for health status at baseline, as in our primary model. The fixed effect for the case/control sampling strata, which are matched on lookback duration, accounts for varying lookback durations across strata. We believe the intervals near the PD index date are potentially biased by confounding by indication while longer intervals from first exposure to diagnosis have less potential confounding, **Figure S1**. Our primary focus in this analysis is on whether the long-run intervals (> 3 years interval) have a stable point estimate and the estimated long run odds ratio.

Third, it is possible that observed association is biased by correlation between the clinical indication for an antipsychotic, which may describe prodromal symptoms of PD, and the use of an antipsychotic. To ensure our estimate describes the marginal effect of the use of an antipsychotic while controlling for the indication, we defined a set of medications plausibly used to treat these symptoms but without clinically meaningful inhibition of D2 to serve as negative controls. We evaluated ever use and duration of exposure of the atypical antidepressant bupropion, the antidepressant and sleep aid trazodone, and the sleep medications eszipcilone, zaleplon, zolpidem, and zopiclone (collectively, “Z drugs”). We estimate the same fixed effects logistic regression as in the primary analysis using these exposures.

Fourth, the medications metoclopramide and ondansetron are both used to manage nausea, vomiting, and gastroparesis and are not subject to the same confounding risks as the antipsychotics from undiagnosed PD. However, metoclopramide exhibits strong D2 affinity (Ki = 64 nM) while ondansetron exhibits functionally no affinity (D2 Ki > 10,000 nM) is a plausible active comparator.

Fifth, in our primary model we control for several factors that could both be symptoms of undiagnosed or prodromal PD and are indications for the use of antipsychotics, potentially introducing collider bias. We seek to address this potential using two analytical frameworks. In the first, we remove the Elixhauser flags for neurological diagnoses and stroke, psychoses, depression, paralysis, and the CCS flags for anxiety (651), delirium and dementia (653), impulse control disorders (656), mood disorders (657) and schizophrenia (659) as potential colliders. This eliminates the conditioning on a collider but exposes us to additional confounding by indication. This motivates our second analysis where we calculate these potential colliders by omitting comorbidities first diagnosed in the 1, 3, or 5 years prior to the PD index date. By applying this time-windowing to the exposure and potential collider calculation, we address both the potential confounding due to undiagnosed PD while also avoiding conditioning on a collider.

Sixth, we conducted further assessment of the potential for confounding by indication. We re-estimated our primary model with an interaction between the ever use indicator with the CCS and Elixhauser flags that potentially represent indications for antipsychotic use (Elixhauser flags for psychoses or depression, CCS flags for anxiety (651), delirium, dementia, and other cognitive disorders (653), impulse control disorders (656), mood disorders (657), personality disorders (658), and schizophrenia (659). For interpretability, we report the average marginal effect – the combination of the main effect for ever use as well as all relevant interactions – of ever use stratified by having any versus none of these diagnoses as well as by the individual diagnostic groups. We estimated the 95% CI for these marginal effect odds ratios using fractional weighting bootstrapping with 9,999 replicates.

Finally, we repeated our analysis individually for the commonly used antipsychotics in our cohort.

All analyses were done using R 4.1.3 (20) using the packages duckdb (21) for data storage, icd for comorbidity calculations (22), fixest for fast fixed effects model estimation (23), and marginaleffects for marginal effects calculation (24).

## Results

After applying our inclusion and exclusion rules, we identified 65,275 newly diagnosed cases of PD and 652,364 matched controls, **Figure S2**. Cohort demographics are described in **Table 2**. During the pre-diagnosis period, relative to controls PD cases had elevated rates of health care utilization, medical complexity, and comorbidities. People who would be diagnosed with PD were more likely to be treated with any antipsychotic (12.74% vs 5.44%) and for longer durations (72.0 vs 17.5 days supplied).

**Table 2:** Sample Summary at Baseline.

|  |  | PD Cases | Non-PD Controls |
| --- | --- | --- | --- |
| Sample Size | N | 65,275 | 652,364 |
| Demographics | Age | 67.5 (11.7) | 67.5 (11.7) |
|  | Female | 25,983 (39.81%) | 359,814 (39.83%) |
|  | Enrollment Start Year | 2007 (5.6) | 2007 (5.6) |
| Health Care Utilization History and Intensity | Inpatient Hospitalizations Per Year | 0.21 (0.40) | 0.15 (0.32) |
|  | Outpatient Encounters Per Year | 18.37 (15.46) | 13.47 (13.25) |
|  | Mean Number of Diagnoses Per Encounter Date | 1.97 (0.95) | 1.83 (0.94) |
| Elixhauser Comorbidities | Alcohol Abuse | 1,358 (2.08%) | 9,924 (1.52%) |
|  | Anemia | 15,306 (23.45%) | 122,692 (18.81%) |
|  | Blood Loss | 1,662 (2.55%) | 14,664 (2.25%) |
|  | Heart Failure | 9,160 (14.03%) | 82,336 (12.62%) |
|  | Coagulopathy | 3,767 (5.77%) | 28,856 (4.42%) |
|  | Diabetes, Without Complications | 18,167 (27.83%) | 166,373 (25.50%) |
|  | Diabetes, With Complications | 8,954 (13.72%) | 74,127 (11.36%) |
|  | Depression | 12,211 (18.71%) | 59,028 (9.05%) |
|  | Drug Abuse | 989 (1.52%) | 5,607 (0.86%) |
|  | Fluid and Electrolyte Disorders | 12,580 (19.27%) | 91,518 (14.03%) |
|  | HIV | 82 (0.13%) | 868 (0.13%) |
|  | Hypertension, Without Complications | 44,886 (68.76%) | 416,233 (63.80%) |
|  | Hypertension, with Complications | 11,375 (17.43%) | 98,981 (15.17%) |
|  | Hypothyroidism | 12,152 (18.62%) | 98,774 (15.14%) |
|  | Liver Disease | 3,106 (4.76%) | 26,873 (4.12%) |
|  | Lymphoma | 1,028 (1.57%) | 9,936 (1.52%) |
|  | Metastatic Cancer | 1,274 (1.95%) | 16,514 (2.53%) |
|  | Neurological Disease | 19,131 (19.31%) | 71,983 (11.03%) |
|  | Obesity | 7,426 (11.38%) | 67,744 (10.38%) |
|  | Pulmonary Hypertension | 2,793 (4.28%) | 23,115 (3.54%) |
|  | Peptic Ulcer Disease | 483 (0.74%) | 4,053 (0.62%) |
|  | Peripheral Vascular Disease | 14,964 (22.92%) | 121,904 (18.69%) |
|  | Paralysis | 3,070 (4.70%) | 13,836 (2.12%) |
|  | Psychoses | 9,810 (15.03%) | 37,934 (5.81%) |
|  | Pulmonary Disease, COPD | 17,656 (27.05%) | 168,168 (25.78%) |
|  | Renal Disease | 5,938 (9.10%) | 54,216 (8.31%) |
|  | Rheumatic Disease | 6,150 (9.42%) | 45,232 (6.93%) |
|  | Tumor | 11,270 (17.27%) | 109,126 (16.73%) |
|  | Valvular Disease | 14,569 (22.32%) | 120,420 (18.46%) |
|  | Weight Loss | 6,051 (9.27%) | 33,983 (5.21%) |
| CCS Codes for Psychiatric Conditions | Anxiety disorders (651) | 11,783 (18.05%) | 63,558 (9.74%) |
|  | Attention-deficit, conduct, and disruptive behavior disorders (652) | 789 (1.21%) | 3,830 (0.59%) |
|  | Delirium, dementia, and amnestic and other cognitive disorders (653) | 11,472 (17.57%) | 40,433 (6.20%) |
|  | Developmental disorders (654) | 595 (0.91%) | 1,707 (0.26%) |
|  | Disorders usually diagnosed in infancy, childhood, or adolescence (655) | 213 (0.33%) | 857 (0.13%) |
|  | Impulse control disorders, NEC (656) | 91 (0.14%) | 424 (0.06%) |
|  | Mood disorders (657) | 15,029 (23.02%) | 69,603 (10.67%) |
|  | Personality disorders (658) | 339 (0.52%) | 1,109 (0.17%) |
|  | Schizophrenia and other psychotic disorders (659) | 3,992 (6.12%) | 13,707 (2.10%) |
|  | Alcohol-related disorders (660) | 1,511 (2.31%) | 11,263 (1.73%) |
|  | Substance-related disorders (661) | 2,042 (3.13%) | 1,038 (0.16%) |
|  | Suicide and intentional self-inflicted injury (662) | 447 (0.68%) | 1,038 (0.16%) |
|  | Screening and history of mental health and substance abuse codes (663) | 5,546 (8.50%) | 57,656 (8.84%) |
|  | Miscellaneous mental health disorders (670) | 4,186 (6.41%) | 23,223 (3.56%) |
| Exposures to Antipsychotics |  |  |  |
| Total Exposure | Ever Use | 8,313 (12.74%) | 35,507 (5.44%) |
|  | *Days Supplied* |  |  |
|  | Average | 72.0 (379.3) | 17.5 (171.8) |
|  | Conditional on Use | 573.0 (926.0) | 325.3 (670.1) |
| Total Exposure Binned By Duration of Use |  |  |  |
|  | Never User | 57,068 (87.4%) | 617,283 (94.6%) |
|  | < 1 Month | 1,404 (2.2%) | 14,303 (2.2%) |
|  | >= 1 Month and < 3 Months | 1,239 (1.9%) | 5,952 (0.9%) |
|  | >= 3 Months and < 6 Months | 879 (1.3%) | 3,000 (0.5%) |
|  | >= 6 Months and < 9 Months | 624 (1.0%) | 1,798 (0.3%) |
|  | >= 9 Months and < 1 Year | 672 (1.0%) | 1,614 (0.2%) |
|  | >= 1 Year and < 2 Years | 1,446 (2.2%) | 3,580 (0.5%) |
|  | >= 2 Years and < 3 Years | 700 (1.1%) | 1,831 (0.3%) |
|  | >= 3 Years | 1,243 (1.9%) | 3,003 (0.5%) |
| By Ki Tier |  |  |  |
| Tier 1: $K_i \leq 1$ nM | Ever Use | 1,832 (2.81%) | 2,991 (0.46%) |
|  | *Days Supplied* |  |  |
|  | Average | 13.17 (127.5) | 1.78 (46.0) |
|  | Conditional on Use | 477.9 (606.8) | 393.1 (560.4) |
| Tier 2: $1 \text{ nM} < K_i < 10 \text{ nM}$ | Ever Use | 4,985 (7.64%) | 25,140 (3.85%) |
|  | *Days Supplied* |  |  |
|  | Average | 27.5 (210.7) | 6.4 (95.1) |
|  | Conditional on Use | 364.2 (682.2) | 168.2 (458.9) |
| Tier 3: $10 \text{ nM} \leq K_i < 100 \text{ nM}$ | Ever Use | 1,964 (3.01%) | 4,918 (0.75%) |
|  | *Days Supplied* |  |  |
|  | Average | 15.8 (157.9) | 3.43 (72.2) |
|  | Conditional on Use | 532.3 (750.3) | 457.9 (698.4) |
| Tier 4: $K_i \geq 100 \text{ nM}$ | Ever Use | 2,441 (3.74%) | 8,268 (1.27%) |
|  | *Days Supplied* |  |  |
|  | Average | 15.5 (151.9) | 5.9 (91.6) |
|  | Conditional on Use | 421.2 (674.7) | 470.2 (672.6) |

Before adjustment, cases had 3.21 (95% CI: 3.17 – 3.26) fold larger odds of ever use compared to controls, **Figure 1**. After adjustment for baseline health utilization and status, this association remained statistically significant (OR = 1.45; 95% CI: 1.40 – 1.51; full model coefficients are reported in **Table S1**). There was a dose-response-like increase in the odds of PD with longer durations of supply, **Figure 1** and **Table S2**.

**Figure 1:**
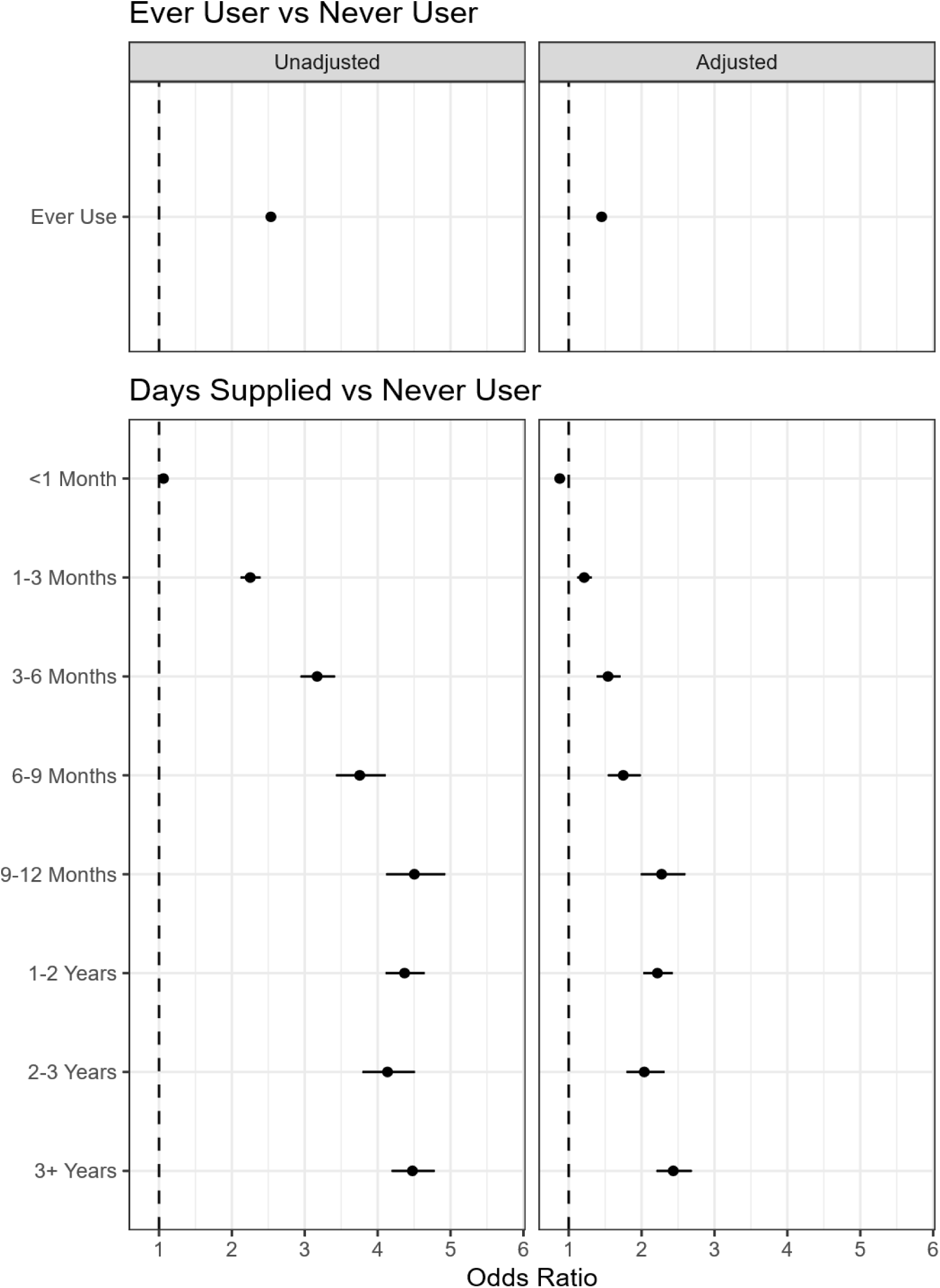
Estimated Associations Between Ever Use, Total Days Supplied, Total Defined Daily Doses Supplied and Development of Parkinson’s Disease. Points reflect the estimated odds ratios while the line range is the 95% confidence interval. Adjusted odds ratios are adjusted for matching (age, sex, time), health care utilization during lookback, and comorbid health conditions at the index date.

The associations varied across the D2 Ki tiers with the greatest adjusted odds ratio with ever use of a compound with a Ki <= 1 nM (OR = 3.53; 95% CI: 3.21 – 3.88) and almost no association with use of the compounds with the lowest affinity (Ki > 100 nM OR = 1.09; 95% CI: 1.02 – 1.17), **Figure 2** and **Table S3**. In the analysis stratifying by duration of exposure, a consistent pattern emerges with the greatest OR for use of the Ki <= 1 nM compounds, similar odds ratios for compounds with Ki values between 1 and 10 nM and between 10 and 100 nM, and almost no association with use of compounds with Ki values greater than 100 nM, **Figure 1** and **Table S4**.

**Figure 2:**
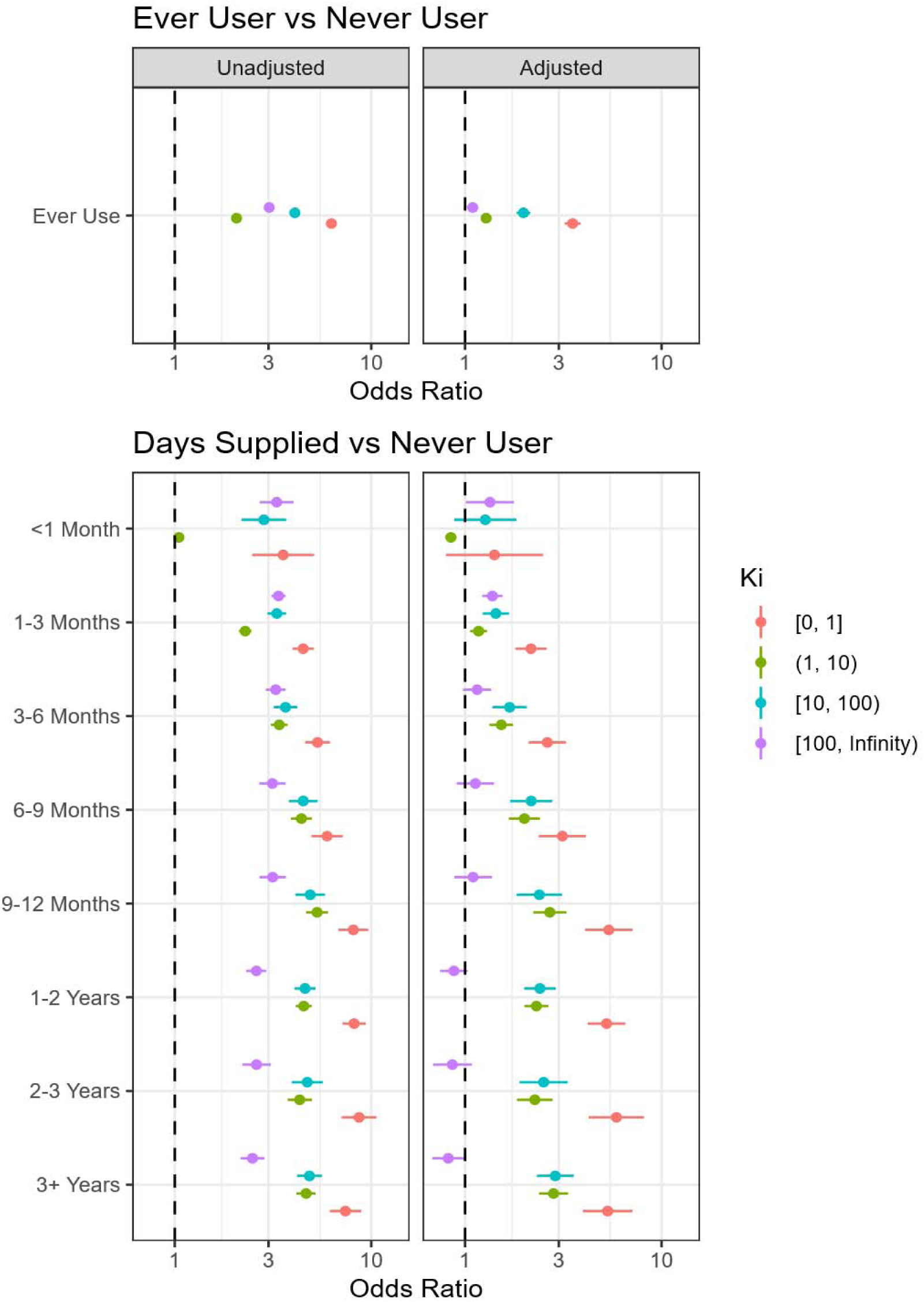
Estimated Associations Between Ever Use, Total Days Supplied and Development of Parkinson’s Disease Stratified By D2 Ki. Points reflect the estimated odds ratios while the line range is the 95% confidence interval. Adjusted odds ratios are adjusted for matching (age, sex, time), health care utilization during lookback, and comorbid health conditions at the index date.

The association between ever use of an antipsychotic and PD is time-varying, **Figure 3**. There is a stronger association between antipsychotic use and diagnosis of PD when the time between exposure is shorter (OR above 1.75 for very short durations before the PD index date); however, this relationship is present at long time horizons (OR = 1.15 for first exposures 4.6-5.7 years prior to the PD diagnosis date). Full model coefficients are reported in **Table S5**.

**Figure 3:**
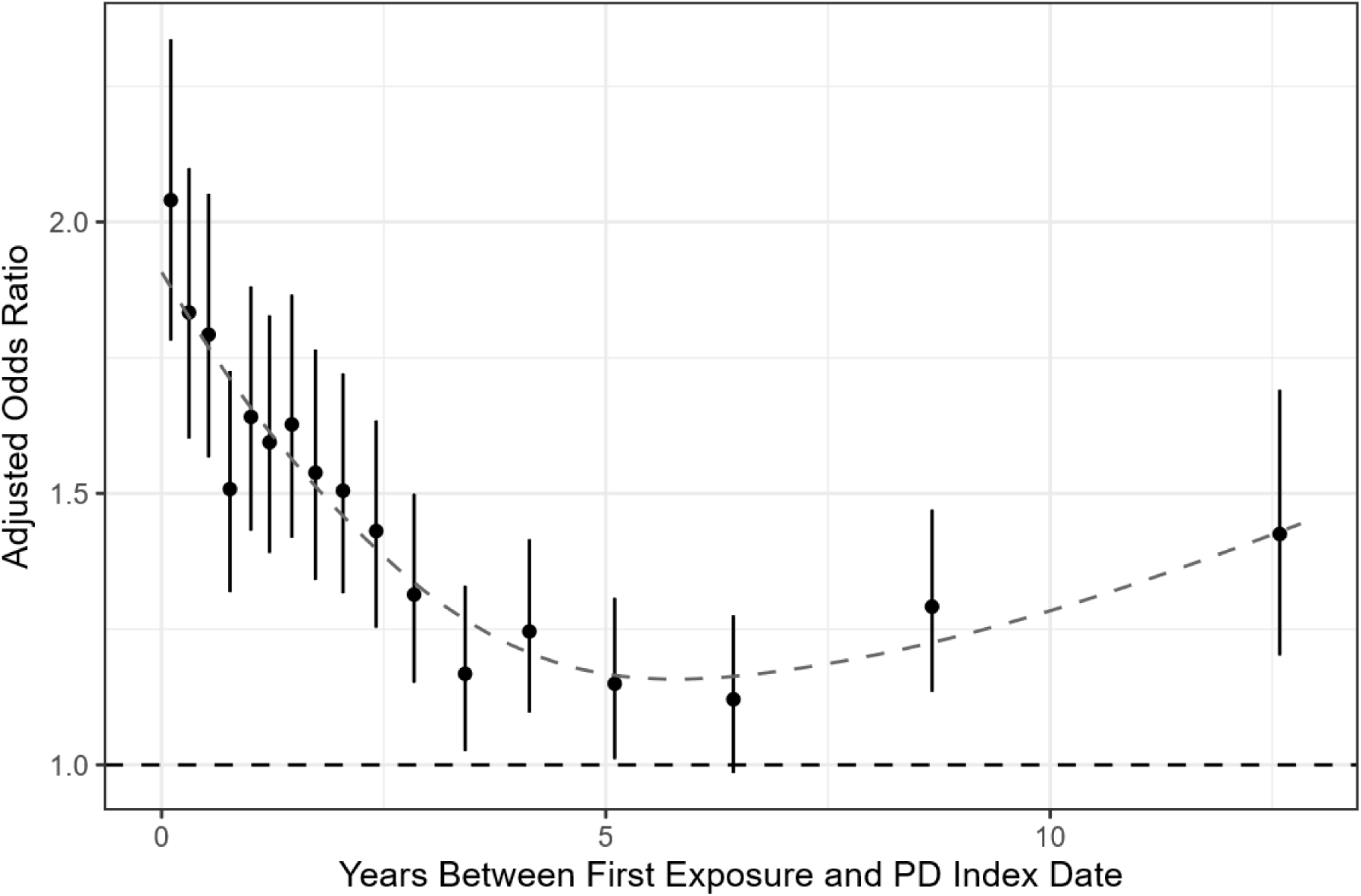
Estimated Associations Between First Use and Development of Parkinson’s Disease With Delayed Follow-Up. We estimated the association between use of antipsychotics and PD by the interval between the first observed dispensing event and the PD index date. Odds ratios are adjusted for matching (age, sex, time), health care utilization during lookback, and comorbid health conditions at the index date. The point reflects the point estimate of the odds ratio while the line range is the 95% confidence interval. Grey dashed line is a generalized additive model smoothed estimate of the overall trend with the points weighted by 1 over the variance of the point estimate.

**Figure 4:**
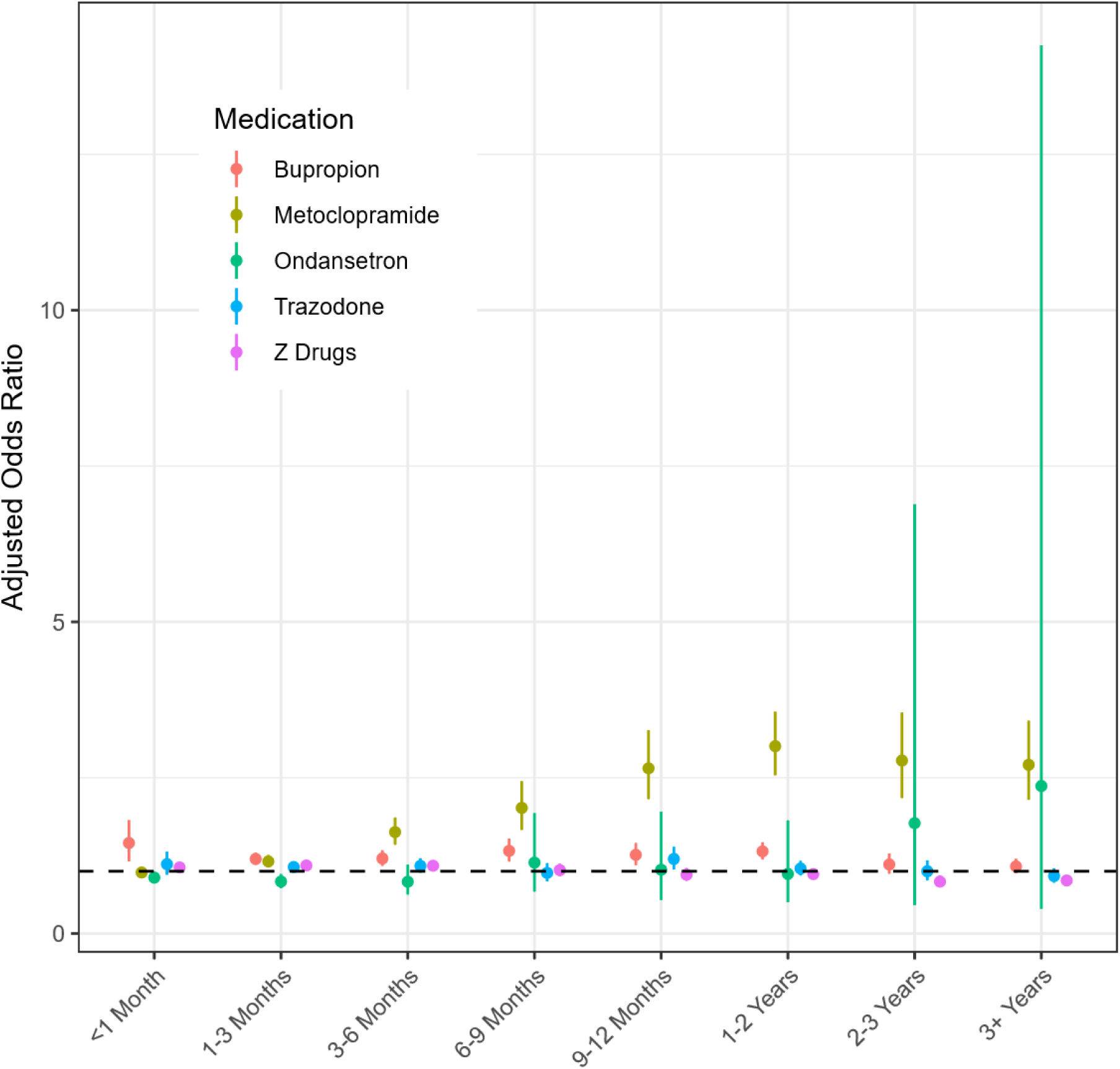
Estimated Associations between Days Supplied and odds of PD for Sensitivity Medications. Points denote the estimated OR and the line range is the 95% CI using standard errors clustered at the level of the sampling group. Odds ratios are compared to never users of the medications and are adjusted for matching (age, sex, time), health care utilization during lookback, and comorbid health conditions at the index date. The very large Cis for ondansetron at long durations are the result of small sample size (n = 25 for 2-3 years, n = 15 for 3+ years).

In our analysis of other medications that may be used to treat psychiatric symptoms or sleep disruptions due to prodromal PD, we found no statistically significant association, after adjustment, between use of trazodone (OR = 1.05; 95% CI: 1.00 – 1.10) or any of the Z-drugs (OR = 1.01; 95% CI: 0.98 – 1.04), **Table S6**. There was an association between ever use of bupropion and PD (OR = 1.21; 95% CI: 1.16 – 1.27). However, none of these medications exhibited an increasing dose-response-like relationship between longer durations of use and PD, **Figure 3** and **Table S7**.

After adjustment, ever use of metoclopramide (Ki = 64) was more common in those with PD (OR = 1.33; 95% CI: 1.27 – 1.39) while use of ondansetron (Ki > 10,000) was largely unassociated with PD (OR = 0.89; 95% CI: 0.85 – 0.94), **Table S6**. The odds of PD was greater in those with longer metoclopramide exposure (OR > 2 for all exposure durations > 6 months) but unchanged in users of ondansetron (OR non-significant after 3 months of use), **Figure 3** and **Table S7**.

Our analyses designed to reveal potential conditioning on a mediator are reported in **Table S8**. In the analysis that removed the potential mediating variables, we observed an increase in the odds associated with ever use (OR = 1.81; 95% CI: 1.74 – 1.87). This increase reflects both new confounding by indication and reduced collider bias. In the time windowed analysis, we saw odds ratios for ever use consistently near 1.55 (OR at 1 year: 1.60 (95% CI: 1.54 – 1.67); at 3 years: 1.52 (95% CI: 1.44 – 1.61), at 5 years: 1.55 (95% CI: 1.44 – 1.67)).

Interaction of psychiatric indications for antipsychotic use with ever use broadly yielded similar estimated odds ratios across all subgroups, **Table S9**. Among patients with any diagnosis suggestive of an indication for antipsychotic use the odds of PD were 1.41 (95% CI: 1.36 – 1.48) greater in users compared to non-users. A similar association was observed among the individuals who never had any diagnosis suggestive of an indication with an odds ratio of 1.53 (95% CI: 1.45 – 1.61), with a similar pattern among the individual diagnostic groups. Among people with an potential indication, use of antipsychotics was associated with greater odds of PD in all for all but one diagnosis group. Only among patients with a diagnosis of delirium or dementia (CCS 653) was the association with exposure to antipsychotics not associated with elevated odds of PD. For patients with an existing dementia diagnosis, it is plausible significant neurodegeneration has already occurred and exposure to antipsychotics has no association due to a saturation effect.

Of the 8 medications with 1,000 or more dispensing events, 7 were associated with greater odds of PD after adjustment, **Table S10**. Only ever use of prochlorperazine was not associated with increased odds of PD. For all medications except quetiapine, longer duration of use is associated with consistently elevated odds of PD, **Table S11**.

## Discussion

We found that ever use of any antipsychotic and, especially, longer durations of use were associated with greater odds of PD. Additionally, we found stronger associations between use of medications with greater affinity for the D2 receptor for the same duration of exposure compared to medications with lower affinity. For antipsychotics with the lowest affinity for D2 receptors (Ki of at least 100 nM), there was no association between greater exposure and odds of PD suggesting the observed associations are driven through the D2 mechanism and not simply by clinical need for an antipsychotic.

The observed estimate (OR = 1.45, E-value = 2.26) may be an underestimate of the true relationship. In the primary analysis, we control for important potential confounders which may also be mediators between the risk of PD and risk of antipsychotic exposure. In analyses designed to reduce this collider bias, we observe increased associations. These increased associations were still present in analyses designed to control for the confounding pathways while avoiding collider bias (OR ∼ 1.55).

The non-monotonic pattern observed for ever use between the Ki-based tiers reflects compositional differences across the tiers. Tier 2 contains prochlorperazine, which was the most common dispensed medication in our sample, which is often used for short durations to treat nausea (median duration of 8 days). This short duration of exposure means the ever use of prochlorperazine represents much less exposure compared to the other medications. However, when stratified by duration, prochlorperazine shows the expected dose-response relationship (**Tables S10-S11**) and the expected ordering of the Ki stratified tiers returns (Tier 1 > Tier 2 ∼ Tier 3 > Tier 4).

Collectively, the results presented here triangulate and support a causal interpretation. Trazodone and the Z-drugs serve as negative controls and the lack of association with PD suggests the observed results are not caused by simple sleep-related prodromal confounding. The metoclopramide results suggest D2 inhibition, not prodromal psychiatric conditions, drives the observed association. The lagged analysis indicates that even newly starting these medications 5 or more years prior to the PD diagnosis date is still associated with increased odds, something unlikely to be caused by prodromal confounding or unmasking of subclinical disease. When the analyses interact use with potential indications for antipsychotics we find elevated odds of PD among exposed patients without psychiatric diagnoses; further evidence implicating the D2 inhibiting medications as the key factor, not unobserved clinical characteristics. Bupropion ever use is associated with greater odds of PD; however, this increase is smaller than that seen with D2 inhibiting medications and lacks the dose-response-like relationship with duration.

This work replicates and extends previous reports with a similar estimated associations conducted in Italy (10) and France (11). Despite long-standing concern about the safety of antipsychotic use in older adults (25), 5.4% of our control enrollees were ever used and approximately 4% were exposed to high-affinity D2 antagonists (Ki < 10 nM). Even lower affinity agents (e.g., second-generation antipsychotics) do not appear to be “safe” in an objective sense.

Intriguingly, the use of these antipsychotics has been increasing over time (26–28), often, as in our cohort, without clear psychiatric indications and by primary care providers (29,30). During the same period, incidence of PD has increased more rapidly than can be explained by age alone. Our data suggest a possible mechanism: increased antipsychotic use may be driving increased incidence of PD. Assuming a causal relationship the use of antipsychotics may explain 2.26% (95% CI: 1.98 – 2.57) of all cases of PD, **Table S12**.

Our analysis faces the limitations inherent to any observational analysis of administrative data. Our key elements – including case, exposure, and confounders – are defined using diagnostic codes of varying accuracy and is limited to the time a person was enrolled in the Marketscan database. Drug-induced or drug-uncovered parkinsonism cannot be fully excluded without clinical detail. We sought to establish a dose-response relationship between a medication’s affinity for D2 measured by the Ki; however, this groups together full and partial antagonists (such as aripiprazole and brexpiprazole). Definitions of the exposure intensity that incorporates more sophisticated pharmacodynamic properties are needed. Future prospective study in humans and in investigator-controlled pre-clinical models is necessary to demonstrate a causal relationship between antipsychotic use and PD.

We consistently found greater odds of PD among people exposed to antipsychotics with an analysis triangulating a potential causative role of this exposure. Antipsychotics are already discouraged in older patients, and our results suggest use may warrant further caution and counseling of risks when prescribing antipsychotics. Our results, if causal, suggest over 2% of all cases of PD may be attributable to – and preventable by avoiding – the use of antipsychotics.

## Data Availability

Merative Marketscan data are used under license from Merative and cannot be redistributed by the authors.

