## Supplemental for "Recent antipsychotics use associated with elevated risk of Parkinson’s disease"

**Figure S1: Theorized Time-Varying Relationship Between Antipsychotic Exposure, PD Risk, and Confounders.** Use of antipsychotics and PD risk may feature confounding, especially when the first use is very recent. Antipsychotics may “unmask” existing disease, either through worsening symptoms or triggering clinical detection leading to a PD diagnosis, or undiagnosed PD may cause psychiatric symptoms that are misdiagnosed and treated with an antipsychotic. However, as the time between the first use increases, these confounding relationships become weaker. Eventually, in the long-run, there is no confounding relationship through the unmasking or treatment of symptoms of undiagnosed PD pathways. The 3 directed acyclic graphs describe the theorized time-varying confounding relationships.

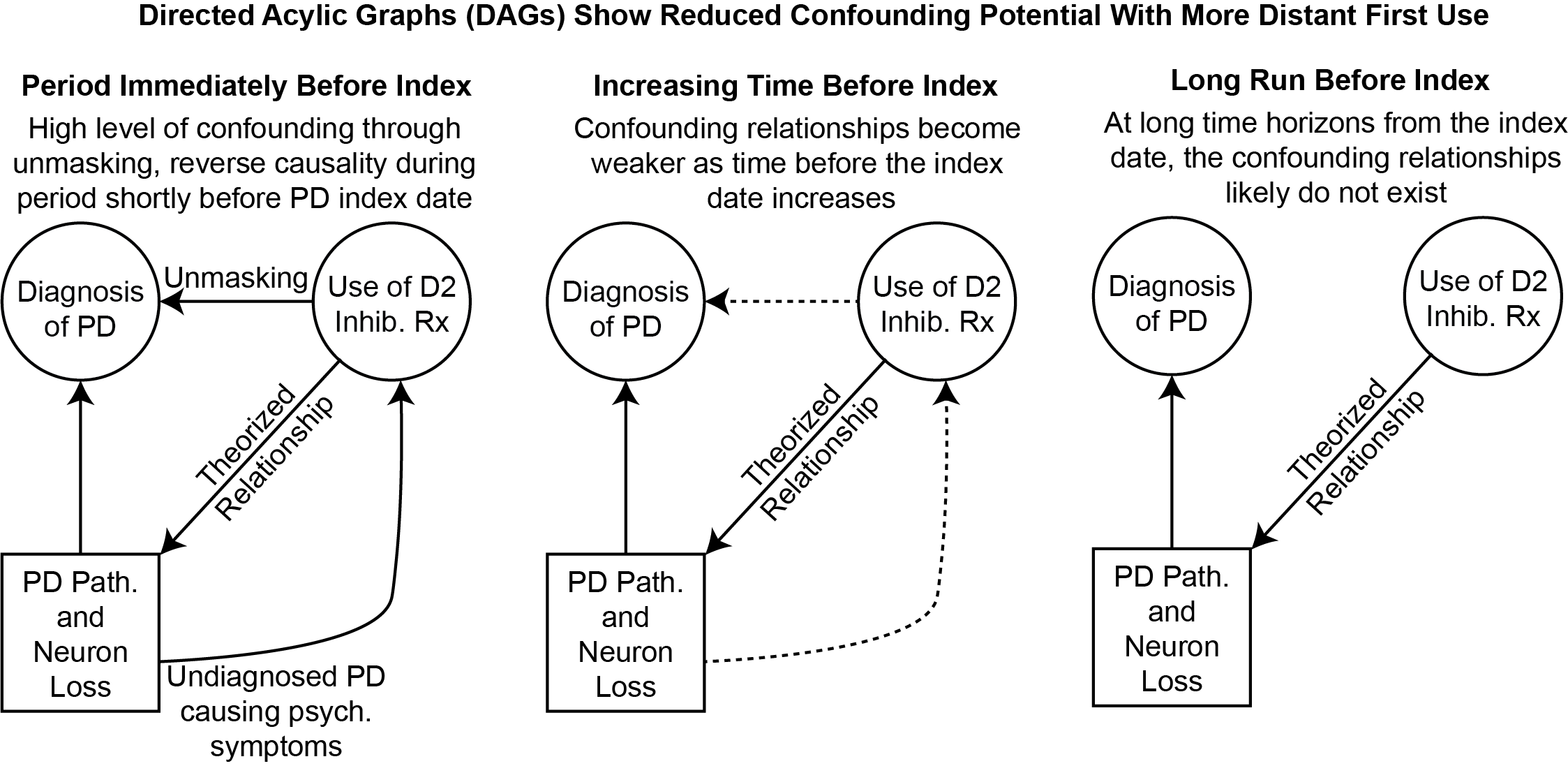

**Figure S2: CONSORT Cohort Construction Flowchart.** Cases were matched exactly on sex and year-of-birth, enrollments of controls were trimmed to exactly match case enrollments. Each case was matched to up to 10 controls. If fewer than 10 controls shared the requires sex, year-of-birth, and enrollment period values to be matched, fewer than 10 controls would be matched to that case. Cases were matched without replacement.

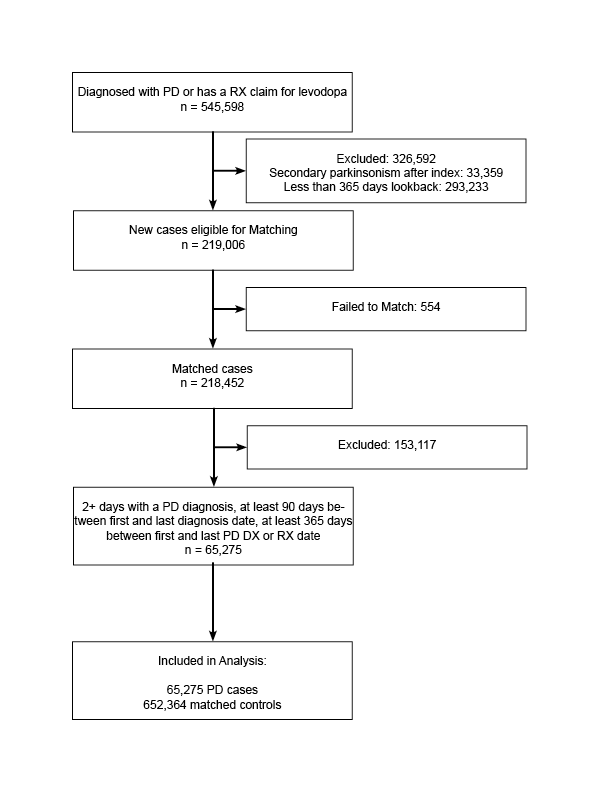

**Table S1: Full Model Estimate for Ever Use of an Antipsychotic.** Model was estimated using a logistic fixed effects regression and included a fixed effects for the sampling strata (1 case and 10 controls matched exactly on age, sex, and enrollment period). As a result of the matching, we do not include age, sex, or time as covariates in the model; the fixed effect for the strata addresses confounding by these variables. Standard errors were clustered by the case/control strata.

|  |  | **95% CI** | |
| --- | --- | --- | --- |
| **Term** | **Odds Ratio** | **Lower Bound** | **Upper Bound** |
| Ever Use | 1.45 | 1.40 | 1.51 |
| Rates of Health Care Utilization During Lookback |  |  |  |
| Inpatient | 0.77 | 0.74 | 0.80 |
| Outpatient | 1.02 | 1.02 | 1.02 |
| Average Number of Diagnoses Per Outpatient Encounter | 1.19 | 1.17 | 1.20 |
| Elixhauser/AHRQ Comorbidities |  |  |  |
| Alcohol Abuse | 0.96 | 0.77 | 1.19 |
| Anemia | 0.99 | 0.96 | 1.02 |
| Blood Loss Amenia | 0.84 | 0.79 | 0.90 |
| Coagulopathy | 0.99 | 0.95 | 1.04 |
| COPD | 0.83 | 0.81 | 0.85 |
| Depression | 1.05 | 0.99 | 1.11 |
| Diabetes without Complications | 0.94 | 0.91 | 0.96 |
| Diabetes with Complications | 1.04 | 1.00 | 1.08 |
| Drug Abuse | 1.12 | 1.00 | 1.27 |
| Fluid and Electrolyte Disorders | 0.98 | 0.95 | 1.01 |
| Heart Failure | 0.77 | 0.74 | 0.80 |
| HIV | 0.67 | 0.52 | 0.88 |
| Hypertension without Complications | 1.08 | 1.05 | 1.10 |
| Hypertension with Complications | 0.98 | 0.95 | 1.01 |
| Hypothyroidism | 1.10 | 1.07 | 1.13 |
| Liver Disorders | 0.93 | 0.88 | 0.97 |
| Lymphoma | 0.82 | 0.75 | 0.89 |
| Metastatic Cancer | 0.52 | 0.49 | 0.56 |
| Neurological Disorders and Stroke | 2.79 | 2.71 | 2.88 |
| Obesity | 0.93 | 0.90 | 0.96 |
| Paralysis | 1.31 | 1.24 | 1.39 |
| Peptic Ulcer Disease | 0.86 | 0.77 | 0.98 |
| Peripheral Vascular Disease | 1.02 | 1.00 | 1.05 |
| Psychoses | 1.28 | 1.21 | 1.35 |
| Pulmonary Hypertension | 0.94 | 0.89 | 0.99 |
| Renal Disorders | 0.77 | 0.74 | 0.81 |
| Rheumatic Disorders | 1.14 | 1.10 | 1.18 |
| Solid Tumor | 0.96 | 0.93 | 0.98 |
| Valvular Disorders | 1.08 | 1.05 | 1.11 |
| Weight Loss | 1.33 | 1.28 | 1.39 |
| Clinical Classification Software Codes |  |  |  |
| Anxiety disorders (651) | 1.39 | 1.35 | 1.44 |
| Attention-deficit, conduct, and disruptive behavior disorders (652) | 0.87 | 0.78 | 0.96 |
| Delirium, dementia, and amnestic and other cognitive disorders (653) | 1.72 | 1.66 | 1.79 |
| Developmental disorders (654) | 1.26 | 1.09 | 1.45 |
| Disorders usually diagnosed in infancy, childhood, or adolescence (655) | 1.45 | 1.18 | 1.79 |
| Impulse control disorders, NEC (656) | 0.65 | 0.47 | 0.88 |
| Mood disorders (657) | 1.46 | 1.37 | 1.56 |
| Personality disorders (658) | 0.96 | 0.81 | 1.14 |
| Schizophrenia and other psychotic disorders (659) | 0.98 | 0.91 | 1.05 |
| Alcohol-related disorders (660) | 0.89 | 0.72 | 1.09 |
| Substance-related disorders (661) | 0.73 | 0.67 | 0.79 |
| Suicide and intentional self-inflicted injury (662) | 1.39 | 1.18 | 1.64 |
| Screening and history of mental health and substance abuse codes (663) | 0.73 | 0.71 | 0.76 |
| Miscellaneous mental health disorders (670) | 1.17 | 1.12 | 1.22 |

**Table S2: Full Model Estimate for Total Days Supplied of All Antipsychotic.** Model was estimated using a logistic fixed effects regression and included a fixed effects for the sampling strata (1 case and 10 controls matched exactly on age, sex, and enrollment period). As a result of the matching, we do not include age, sex, or time as covariates in the model; the fixed effect for the strata addresses confounding by these variables. Standard errors were clustered by the case/control strata.

|  |  | **95% CI** | |
| --- | --- | --- | --- |
| **Term** | **Odds Ratio** | **Lower Bound** | **Upper Bound** |
| Days Supplied |  |  |  |
| Never User | Reference | | |
| < 1 Month | 0.88 | 0.82 | 0.94 |
| >= 1 Month and < 3 Months | 1.21 | 1.12 | 1.32 |
| >= 3 Months and < 6 Months | 1.54 | 1.38 | 1.71 |
| >= 6 Months and < 9 Months | 1.75 | 1.53 | 1.99 |
| >= 9 Months and < 1 Year | 2.27 | 1.99 | 2.60 |
| >= 1 Year and < 2 Years | 2.22 | 2.02 | 2.43 |
| >= 2 Years and < 3 Years | 2.04 | 1.79 | 2.32 |
| >= 3 Years | 2.43 | 2.20 | 2.69 |
| Rates of Health Care Utilization During Lookback |  |  |  |
| Inpatient | 0.77 | 0.74 | 0.80 |
| Outpatient | 1.02 | 1.02 | 1.02 |
| Average Number of Diagnoses Per Outpatient Encounter | 1.19 | 1.18 | 1.21 |
| Elixhauser/AHRQ Comorbidities |  |  |  |
| Alcohol Abuse | 0.95 | 0.76 | 1.19 |
| Anemia | 0.99 | 0.97 | 1.02 |
| Blood Loss Amenia | 0.84 | 0.79 | 0.90 |
| Coagulopathy | 1.00 | 0.95 | 1.05 |
| COPD | 0.83 | 0.81 | 0.85 |
| Depression | 1.06 | 1.00 | 1.13 |
| Diabetes without Complications | 0.94 | 0.91 | 0.96 |
| Diabetes with Complications | 1.04 | 1.00 | 1.08 |
| Drug Abuse | 1.11 | 0.98 | 1.25 |
| Fluid and Electrolyte Disorders | 0.99 | 0.96 | 1.02 |
| Heart Failure | 0.77 | 0.74 | 0.79 |
| HIV | 0.67 | 0.51 | 0.87 |
| Hypertension without Complications | 1.08 | 1.05 | 1.10 |
| Hypertension with Complications | 0.98 | 0.95 | 1.01 |
| Hypothyroidism | 1.09 | 1.06 | 1.12 |
| Liver Disorders | 0.93 | 0.89 | 0.98 |
| Lymphoma | 0.86 | 0.80 | 0.93 |
| Metastatic Cancer | 0.57 | 0.53 | 0.61 |
| Neurological Disorders and Stroke | 2.78 | 2.70 | 2.87 |
| Obesity | 0.93 | 0.89 | 0.96 |
| Paralysis | 1.31 | 1.24 | 1.39 |
| Peptic Ulcer Disease | 0.87 | 0.77 | 0.98 |
| Peripheral Vascular Disease | 1.02 | 1.00 | 1.05 |
| Psychoses | 1.23 | 1.16 | 1.30 |
| Pulmonary Hypertension | 0.95 | 0.90 | 1.00 |
| Renal Disorders | 0.77 | 0.74 | 0.80 |
| Rheumatic Disorders | 1.15 | 1.11 | 1.19 |
| Solid Tumor | 0.97 | 0.94 | 1.00 |
| Valvular Disorders | 1.09 | 1.06 | 1.11 |
| Weight Loss | 1.34 | 1.28 | 1.39 |
| Clinical Classification Software Codes |  |  |  |
| Anxiety disorders (651) | 1.39 | 1.34 | 1.43 |
| Attention-deficit, conduct, and disruptive behavior disorders (652) | 0.85 | 0.76 | 0.94 |
| Delirium, dementia, and amnestic and other cognitive disorders (653) | 1.68 | 1.61 | 1.74 |
| Developmental disorders (654) | 1.20 | 1.04 | 1.38 |
| Disorders usually diagnosed in infancy, childhood, or adolescence (655) | 1.42 | 1.15 | 1.75 |
| Impulse control disorders, NEC (656) | 0.62 | 0.45 | 0.85 |
| Mood disorders (657) | 1.42 | 1.33 | 1.52 |
| Personality disorders (658) | 0.88 | 0.74 | 1.05 |
| Schizophrenia and other psychotic disorders (659) | 0.94 | 0.87 | 1.01 |
| Alcohol-related disorders (660) | 0.89 | 0.72 | 1.09 |
| Substance-related disorders (661) | 0.72 | 0.67 | 0.78 |
| Suicide and intentional self-inflicted injury (662) | 1.30 | 1.10 | 1.53 |
| Screening and history of mental health and substance abuse codes (663) | 0.73 | 0.71 | 0.76 |
| Miscellaneous mental health disorders (670) | 1.17 | 1.11 | 1.22 |

**Table S3: Full Model Estimate for Ever Use of an Antipsychotic by D2 Ki Tier.** Model was estimated using a logistic fixed effects regression and included a fixed effects for the sampling strata (1 case and 10 controls matched exactly on age, sex, and enrollment period). As a result of the matching, we do not include age, sex, or time as covariates in the model; the fixed effect for the strata addresses confounding by these variables. Standard errors were clustered by the case/control strata.

| **Term** | **Ki <= 1 nM** | **1 < Ki < 10 nM** | **10 <= Ki < 100 nM** | **Ki >= 100 nM** |
| --- | --- | --- | --- | --- |
| Ever Use | 3.53  (3.21, 3.88) | 1.28  (1.22, 1.34) | 1.98  (1.83, 2.15) | 1.09  (1.02, 1.17) |
| Rates of Health Care Utilization During Lookback |  |  |  |  |
| Inpatient | 0.78  (0.75, 0.81) | 0.77  (0.75, 0.80) | 0.77  (0.75, 0.80) | 0.78  (0.75, 0.81) |
| Outpatient | 1.02  (1.02, 1.02) | 1.02  (1.02, 1.02) | 1.02  (1.02, 1.02) | 1.02  (1.02, 1.02) |
| Average Number of Diagnoses Per Outpatient Encounter | 1.19  (1.18, 1.21) | 1.19  (1.17, 1.20) | 1.19  (1.17, 1.20) | 1.18  (1.17, 1.20) |
| Elixhauser/AHRQ Comorbidities |  |  |  |  |
| Alcohol Abuse | 0.95  (0.76, 1.18) | 0.97  (0.78, 1.21) | 0.96  (0.77, 1.20) | 0.97  (0.77, 1.20) |
| Anemia | 0.99  (0.97, 1.02) | 0.99  (0.96, 1.02) | 0.99  (0.97, 1.02) | 0.99  (0.97, 1.02) |
| Blood Loss Amenia | 0.84  (0.79, 0.90) | 0.84  (0.79, 0.90) | 0.84  (0.79, 0.90) | 0.84  (0.78, 0.90) |
| Coagulopathy | 0.99  (0.95, 1.04) | 0.99  (0.95, 1.04) | 0.99  (0.95, 1.04) | 0.99  (0.95, 1.04) |
| COPD | 0.83  (0.81, 0.85) | 0.83  (0.81, 0.85) | 0.83  (0.81, 0.85) | 0.83  (0.81, 0.85) |
| Depression | 1.01  (0.95, 1.07) | 1.05  (0.99, 1.11) | 1.05  (0.99, 1.11) | 1.04  (0.98, 1.10) |
| Diabetes without Complications | 0.94  (0.91, 0.96) | 0.94  (0.92, 0.97) | 0.94  (0.92, 0.97) | 0.94  (0.92, 0.97) |
| Diabetes with Complications | 1.04  (1.00, 1.07) | 1.04  (1.00, 1.07) | 1.04  (1.00, 1.08) | 1.03  (1.00, 1.07) |
| Drug Abuse | 1.12  (0.99, 1.26) | 1.14  (1.02, 1.29) | 1.13  (1.01, 1.27) | 1.14  (1.01, 1.28) |
| Fluid and Electrolyte Disorders | 0.99  (0.96, 1.02) | 0.99  (0.96, 1.02) | 0.99  (0.96, 1.02) | 0.99  (0.96, 1.02) |
| Heart Failure | 0.77  (0.74, 0.80) | 0.77  (0.74, 0.80) | 0.77  (0.74. 0.80) | 0.77  (0.74, 0.80) |
| HIV | 0.69  (0.53, 0.90) | 0.67  (0.52, 0.88) | 0.67  (0.52, 0.88) | 0.68  (0.52, 0.88) |
| Hypertension without Complications | 1.08  (1.05, 1.10) | 1.08  (1.05, 1.10) | 1.08  (1.05, 1.10) | 1.08  (1.05, 1.10) |
| Hypertension with Complications | 0.98  (0.95, 1.01) | 0.98  (0.95, 1.01) | 0.98  (0.95, 1.01) | 0.98  (0.95, 1.01) |
| Hypothyroidism | 1.09  (1.06, 1.12) | 1.10  (1.07, 1.13) | 1.10  (1.07, 1.13) | 1.10  (1.07, 1.13) |
| Liver Disorders | 0.93  (0.89, 0.98) | 0.93  (0.88, 0.97) | 0.93  (0.89, 0.98) | 0.93  (0.89, 0.98) |
| Lymphoma | 0.86  (0.79, 0.93) | 0.83  (0.76, 0.90) | 0.85  (0.79, 0.92) | 0.85  (0.79, 0.92) |
| Metastatic Cancer | 0.56  (0.52, 0.61) | 0.54  (0.50, 0.58) | 0.56  (0.52, 0.60) | 0.56  (0.52, 0.60) |
| Neurological Disorders and Stroke | 2.82  (2.73, 2.90) | 2.81  (2.73, 2.90) | 2.81  (2.73, 2.90) | 2.82  (2.73, 2.91) |
| Obesity | 0.92  (0.89, 0.95) | 0.92  (0.89, 0.96) | 0.93  (0.90, 0.96) | 0.92  (0.89, 0.96) |
| Paralysis | 1.31  (1.24, 1.39) | 1.31  (1.23, 1.38) | 1.31  (1.23, 1.38) | 1.30  (1.23, 1.38) |
| Peptic Ulcer Disease | 0.86  (0.76, 0.97) | 0.86  (0.77, 0.97) | 0.87  (0.77, 0.98) | 0.87  (0.77, 0.98) |
| Peripheral Vascular Disease | 1.03  (1.00, 1.05) | 1.02  (1.00, 1.05) | 1.02  (1.00, 1.05) | 1.02  (1.00, 1.05) |
| Psychoses | 1.23  (1.16, 1.30) | 1.33  (1.26, 1.41) | 1.32  (1.24, 1.39) | 1.34  (1.27, 1.42) |
| Pulmonary Hypertension | 0.94  (0.89, 0.99) | 0.94  (0.89, 0.99) | 0.94  (0.89, 0.99) | 0.94  (0.89, 0.99) |
| Renal Disorders | 0.77  (0.74, 0.80) | 0.77  (0.74, 0.80) | 0.77  (0.74, 0.80) | 0.77  (0.74, 0.80) |
| Rheumatic Disorders | 1.14  (1.10, 1.18) | 1.14  (1.10, 1.18) | 1.14  (1.10, 1.18) | 1.14  (1.10, 1.18) |
| Solid Tumor | 0.97  (0.94, 0.99) | 0.96  (0.93, 0.99) | 0.97  (0.94, 0.99) | 0.96  (0.94, 0.99) |
| Valvular Disorders | 1.08  (1.05, 1.11) | 1.08  (1.05, 1.11) | 1.08  (1.05, 1.11) | 1.08  (1.05, 1.11) |
| Weight Loss | 1.34  (1.29, 1.40) | 1.34  (1.29, 1.39) | 1.34  (1.28, 1.39) | 1.34  (1.29, 1.40) |
| Clinical Classification Software Codes |  |  |  |  |
| Anxiety disorders (651) | 1.39  (1.35, 1.44) | 1.41  (1.36, 1.45) | 1.41  (1.36, 1.45) | 1.41  (1.37, 1.46) |
| Attention-deficit, conduct, and disruptive behavior disorders (652) | 0.84  (0.75, 0.93) | 0.88  (0.80, 0.98) | 0.88  (0.79, 0.98) | 0.89  (0.80, 0.99) |
| Delirium, dementia, and amnestic and other cognitive disorders (653) | 1.80  (1.73, 1.87) | 1.77  (1.71, 1.84) | 1.76  (1.70, 1.83) | 1.79  (1.73, 1.86) |
| Developmental disorders (654) | 1.30  (1.13, 1.50) | 1.29  (1.12, 1.49) | 1.29  (1.12, 1.48) | 1.31  (1.14, 1.51) |
| Disorders usually diagnosed in infancy, childhood, or adolescence (655) | 1.41  (1.14, 1.74) | 1.46  (1.18, 1.79) | 1.47  (1.19, 1.81) | 1.47  (1.19, 1.81) |
| Impulse control disorders, NEC (656) | 0.66  (0.49, 0.91) | 0.66  (0.48, 0.90) | 0.67  (0.49, 0.91) | 0.67  (0.49, 0.92) |
| Mood disorders (657) | 1.51  (1.41, 1.61) | 1.48  (1.39, 1.58) | 1.47  (1.38, 1.57) | 1.50  (1.40, 1.60) |
| Personality disorders (658) | 0.93  (0.78, 1.11) | 0.99  (0.83, 1.17) | 0.96  (0.81, 1.15) | 1.01  (0.85, 1.20) |
| Schizophrenia and other psychotic disorders (659) | 1.03  (0.96, 1.11) | 0.99  (0.92, 1.06) | 0.98  (0.91, 1.05) | 1.01  (0.94, 1.09) |
| Alcohol-related disorders (660) | 0.89  (0.72, 1.10) | 0.88  (0.72, 1.09) | 0.88  (0.72, 1.09) | 0.88  (0.71, 1.08) |
| Substance-related disorders (661) | 0.72  (0.67, 0.78) | 0.73  (0.68, 0.79) | 0.73  (0.67, 0.79) | 0.73  (0.68, 0.79) |
| Suicide and intentional self-inflicted injury (662) | 1.21  (1.02, 1.44) | 1.46  (1.24, 1.72) | 1.40  (1.19, 1.65) | 1.47  (1.25, 1.73) |
| Screening and history of mental health and substance abuse codes (663) | 0.73  (0.71, 0.76) | 0.73  (0.71, 0.76) | 0.73  (0.71, 0.76) | 0.73  (0.71, 0.76) |
| Miscellaneous mental health disorders (670) | 1.17  (1.11, 1.22) | 1.17  (1.12, 1.23) | 1.17  (1.12, 1.23) | 1.17  (1.12, 1.23) |

**Table S4: Full Model Estimate for Total Days Supplied of All Antipsychotic by D2 Ki Tier.** Model was estimated using a logistic fixed effects regression and included a fixed effects for the sampling strata (1 case and 10 controls matched exactly on age, sex, and enrollment period). As a result of the matching, we do not include age, sex, or time as covariates in the model; the fixed effect for the strata addresses confounding by these variables. Standard errors were clustered by the case/control strata.

| **Term** | **Ki <= 1 nM** | **1 < Ki < 10 nM** | **10 <= Ki < 100 nM** | **Ki >= 100 nM** |
| --- | --- | --- | --- | --- |
| Days Supplied |  |  |  |  |
| Never User | Reference | Reference | Reference | Reference |
| < 1 Month | 1.41  (0.80, 2.50) | 0.85  (0.79, 0.91) | 1.27  (0.88, 1.83) | 1.34  (1.01, 1.78) |
| >= 1 Month and < 3 Months | 2.17  (1.80, 2.60) | 1.17  (1.06, 1.29) | 1.43  (1.23, 1.68) | 1.38  (1.22, 1.55) |
| >= 3 Months and < 6 Months | 2.62  (2.11, 3.27) | 1.53  (1.33, 1.76) | 1.69  (1.38, 2.06) | 1.15  (0.98, 1.36) |
| >= 6 Months and < 9 Months | 3.13  (2.37, 4.13) | 2.01  (1.67, 2.41) | 2.17  (1.69, 2.78) | 1.13  (0.91, 1.41) |
| >= 9 Months and < 1 Year | 5.39  (4.07, 7.13) | 2.70  (2.22, 3.28) | 2.39  (1.83, 3.12) | 1.10  (0.88, 1.37) |
| >= 1 Year and < 2 Years | 5.25  (4.21, 6.55) | 2.31  (2.00, 2.66) | 2.41  (2.00, 2.90) | 0.88  (0.75, 1.03) |
| >= 2 Years and < 3 Years | 5.89  (4.26, 8.15) | 2.26  (1.84, 2.79) | 2.51  (1.89, 3.34) | 0.86  (0.69, 1.08) |
| >= 3 Years | 5.32  (3.97, 7.11) | 2.82  (2.38, 3.35) | 2.88  (2.32, 3.58) | 0.82  (0.68, 0.99) |
| Rates of Health Care Utilization During Lookback |  |  |  |  |
| Inpatient | 0.78  (0.75, 0.81) | 0.77  (0.75, 0.80) | 0.78  (0.75, 0.80) | 0.78  (0.75, 0.81) |
| Outpatient | 1.02  (1.02, 1.02) | 1.02  (1.02, 1.02) | 1.02  (1.02, 1.02) | 1.02  (1.02, 1.02) |
| Average Number of Diagnoses Per Outpatient Encounter | 1.19  (1.18, 1.21) | 1.19  (1.17, 1.20) | 1.19  (1.17, 1.20) | 1.18  (1.17, 1.20) |
| Elixhauser/AHRQ Comorbidities |  |  |  |  |
| Alcohol Abuse | 0.95  (0.76, 1.18) | 0.97  (0.78, 1.21) | 0.96  (0.77, 1.20) | 0.97  (0.78, 1.21) |
| Anemia | 0.99  (0.97, 1.02) | 0.99  (0.97, 1.02) | 0.99  (0.97, 1.02) | 0.99  (0.97, 1.02) |
| Blood Loss Amenia | 0.94  (0.79, 0.90) | 0.84  (0.79, 0.90) | 0.84 (0.78, 0.90) | 0.84  (0.78, 0.90) |
| Coagulopathy | 0.99  (0.95, 1.04) | 1.00  (0.95, 1.04) | 0.99  (0.95, 1.04) | 0.99  (0.95, 1.04) |
| COPD | 0.83  (0.81, 0.85) | 0.83  (0.81, 0.85) | 0.83  (0.81, 0.85) | 0.83  (0.81, 0.85) |
| Depression | 1.01  (0.95, 1.07) | 1.07  (1.01, 1.13) | 1.05  (0.99, 1.12) | 1.04  (0.98, 1.10) |
| Diabetes without Complications | 0.94  (0.91, 0.96) | 0.94  (0.91, 0.96) | 0.94  (0.91, 0.96) | 0.94  (0.92, 0.97) |
| Diabetes with Complications | 1.03  (1.00, 1.07) | 1.04  (1.00, 1.07) | 1.04  (1.00, 1.08) | 1.03  (1.00, 1.07) |
| Drug Abuse | 1.12  (0.99, 1.26) | 1.14  (1.01, 1.28) | 1.13  (1.01, 1.27) | 1.14  (1.01, 1.28) |
| Fluid and Electrolyte Disorders | 0.99  (0.96, 1.03) | 0.99  (0.96, 1.02) | 0.99  (0.96, 1.02) | 0.99  (0.96, 1.02) |
| Heart Failure | 0.77  (0.74, 0.80) | 0.77  (0.74, 0.79) | 0.77  (0.74, 0.80) | 0.77  (0.74, 0.80) |
| HIV | 0.69  (0.53, 0.90) | 0.67  (0.51, 0.88) | 0.67  (0.51, 0.88) | 0.68  (0.52, 0.89) |
| Hypertension without Complications | 1.08  (1.05, 1.10) | 1.08  (1.05, 1.10) | 1.08  (1.05, 1.10) | 1.08  (1.05, 1.10) |
| Hypertension with Complications | 0.98  (0.95, 1.01) | 0.98  (0.95, 1.01) | 0.98  (0.95, 1.01) | 0.98  (0.95, 1.01) |
| Hypothyroidism | 1.09  (1.06, 1.12) | 1.10  (1.07, 1.13) | 1.10  (1.07, 1.13) | 1.10  (1.07, 1.13) |
| Liver Disorders | 0.93  (0.89, 0.98) | 0.93  (0.89, 0.98) | 0.93  (0.89, 0.98) | 0.93  (0.89, 0.98) |
| Lymphoma | 0.86  (0.79, 0.93) | 0.86  (0.79, 0.93) | 0.85  (0.79, 0.92) | 0.85  (0.79, 0.92) |
| Metastatic Cancer | 0.56  (0.53, 0.61) | 0.57  (0.53, 0.62) | 0.56  (0.52, 0.60) | 0.56  (0.52, 0.61) |
| Neurological Disorders and Stroke | 2.82  (2.73, 2.90) | 2.81  (2.72, 2.89) | 2.82  (2.73, 2.90) | 2.82  (2.73, 2.91) |
| Obesity | 0.92  (0.89, 0.95) | 0.92  (0.89, 0.96) | 0.93  (0.90, 0.96) | 0.92  (0.89, 0.96) |
| Paralysis | 1.31  (1.24, 1.39) | 1.31  (1.23, 1.39) | 1.31  (1.23, 1.39) | 1.30  (1.23, 1.38) |
| Peptic Ulcer Disease | 0.86  (0.76, 0.97) | 0.86  (0.77, 0.97) | 0.87  (0.77, 0.98) | 0.86  (0.77, 0.97) |
| Peripheral Vascular Disease | 1.02  (1.00, 1.05) | 1.02  (1.00, 1.05) | 1.02  (1.00, 1.05) | 1.02  (1.00, 1.05) |
| Psychoses | 1.23  (1.16, 1.30) | 1.32  (1.25, 1.39) | 1.31  (1.24, 1.39) | 1.34  (1.27, 1.42) |
| Pulmonary Hypertension | 0.94  (0.89, 0.99) | 0.94  (0.89, 0.99) | 0.94  (0.89, 0.99) | 0.94  (0.89, 0.99) |
| Renal Disorders | 0.78  (0.74, 0.80) | 0.77  (0.74, 0.80) | 0.77  (0.74, 0.80) | 0.78  (0.74, 0.80) |
| Rheumatic Disorders | 1.14  (1.10, 1.18) | 1.14  (1.10, 1.19) | 1.14  (1.10, 1.18) | 1.14  (1.10, 1.18) |
| Solid Tumor | 0.97  (0.94, 0.99) | 0.97 (0.94, 1.00) | 0.97  (0.94, 0.99) | 0.96  (0.94, 0.99) |
| Valvular Disorders | 1.08  (1.05, 1.11) | 1.08  (1.05, 1.11) | 1.08  (1.05, 1.11) | 1.08  (1.05, 1.11) |
| Weight Loss | 1.34  (1.29, 1.40) | 1.34  (1.29, 1.40) | 1.34  (1.29, 1.39) | 1.34  (1.29, 1.40) |
| Clinical Classification Software Codes |  |  |  |  |
| Anxiety disorders (651) | 1.40  (1.35, 1.44) | 1.41  (1.36, 1.45) | 1.41  (1.36, 1.45) | 1.41  (1.37, 1.46) |
| Attention-deficit, conduct, and disruptive behavior disorders (652) | 0.84  (0.75, 0.93) | 0.88  (0.79, 0.98) | 0.89  (0.80, 0.98) | 0.89  (0.80, 0.99) |
| Delirium, dementia, and amnestic and other cognitive disorders (653) | 1.80  (1.74, 1.88) | 1.74  (1.67, 1.80) | 1.77  (1.70, 1.84) | 1.80  (1.73, 1.87) |
| Developmental disorders (654) | 1.30  (1.13, 1.50) | 1.25  (1.08, 1.44) | 1.29  (1.13, 1.49) | 1.32  (1.14, 1.52) |
| Disorders usually diagnosed in infancy, childhood, or adolescence (655) | 1.41  (1.14, 1.74) | 1.43  (1.16, 1.76) | 1.47  (1.19, 1.81) | 1.48  (1.20, 1.82) |
| Impulse control disorders, NEC (656) | 0.66  (0.49, 0.91) | 0.63  (0.46, 0.86) | 0.68  (0.49, 0.92) | 0.68  (0.50, 0.92) |
| Mood disorders (657) | 1.51  (1.41, 1.61) | 1.44  (1.35, 1.54) | 1.46  (1.37, 1.56) | 1.50  (1.41, 1.60) |
| Personality disorders (658) | 0.93  (0.79, 1.11) | 0.93  (0.78, 1.11) | 0.97  (0.81, 1.15) | 1.02  (0.86, 1.21) |
| Schizophrenia and other psychotic disorders (659) | 1.04  (0.97, 1.11) | 0.94  (0.87, 1.01) | 0.98  (0.91, 1.05) | 1.02  (0.95, 1.09) |
| Alcohol-related disorders (660) | 0.89  (0.72, 1.10) | 0.88  (0.71, 1.08) | 0.88  (0.72, 1.09) | 0.88  (0.71, 1.08) |
| Substance-related disorders (661) | 0.72  (0.67, 0.78) | 0.73  (0.67, 0.79) | 0.73  (0.67, 0.79) | 0.73  (0.68, 0.80) |
| Suicide and intentional self-inflicted injury (662) | 1.23  (1.04, 1.46) | 1.40  (1.19, 1.66) | 1.41  (1.19, 1.66) | 1.48  (1.26, 1.75) |
| Screening and history of mental health and substance abuse codes (663) | 0.73  (0.70, 0.76) | 0.73  (0.71, 0.76) | 0.73  (0.71, 0.76) | 0.73  (0.71, 0.76) |
| Miscellaneous mental health disorders (670) | 1.17  (1.12, 1.22) | 1.17  (1.12, 1.23) | 1.17  (1.12, 1.23) | 1.17  (1.12, 1.23) |

**Table S5: Full Model Estimate for Ever Use Stratified by Interval Between First Use and PD Index Date.** Model was estimated using a logistic fixed effects regression and included a fixed effects for the sampling strata (1 case and 10 controls matched exactly on age, sex, and enrollment period). As a result of the matching, we do not include age, sex, or time as covariates in the model; the fixed effect for the strata addresses confounding by these variables. Standard errors were clustered by the case/control strata.

|  | | | | |  | **95% CI** | |
| --- | --- | --- | --- | --- | --- | --- | --- |
| **Term** | | | | | **Odds Ratio** | **Lower Bound** | **Upper Bound** |
| Interval Between First Use and PD Diagnosis Date (Years) | | | | |  |  |  |
| Time Period | Start Time | End Time | Number Treated | |  |  |  |
|  | | | Cases | Controls |  |  |  |
| Never User | | | 56,962 | 616,857 | Reference | | |
| 1 | 0.00 | 0.20 | 498 | 1,792 | 2.04 | 1.78 | 2.34 |
| 2 | 0.21 | 0.42 | 497 | 1,867 | 1.83 | 1.60 | 2.10 |
| 3 | 0.42 | 0.64 | 503 | 1,793 | 1.79 | 1.57 | 2.05 |
| 4 | 0.65 | 0.90 | 498 | 1,977 | 1.51 | 1.32 | 1.73 |
| 5 | 0.90 | 1.10 | 501 | 1,889 | 1.64 | 1.43 | 1.88 |
| 6 | 1.11 | 1.33 | 500 | 1,878 | 1.59 | 1.39 | 1.83 |
| 7 | 1.33 | 1.59 | 494 | 1,891 | 1.63 | 1.42 | 1.87 |
| 8 | 1.59 | 1.88 | 498 | 1,960 | 1.54 | 1.34 | 1.77 |
| 9 | 1.88 | 2.22 | 509 | 2,015 | 1.50 | 1.32 | 1.72 |
| 10 | 2.22 | 2.61 | 499 | 2,097 | 1.43 | 1.25 | 1.63 |
| 11 | 2.62 | 3.10 | 501 | 2,214 | 1.31 | 1.15 | 1.50 |
| 12 | 3.11 | 3.76 | 501 | 2,449 | 1.17 | 1.02 | 1.33 |
| 13 | 3.76 | 4.56 | 499 | 2,409 | 1.25 | 1.10 | 1.42 |
| 14 | 4.57 | 5.66 | 501 | 2,586 | 1.15 | 1.01 | 1.31 |
| 15 | 5.66 | 7.35 | 500 | 2,724 | 1.12 | 0.98 | 1.28 |
| 16 | 7.35 | 10.37 | 500 | 2,516 | 1.29 | 1.13 | 1.47 |
| 17 | 10.37 | --- | 314 | 1,450 | 1.43 | 1.20 | 1.69 |
| Rates of Health Care Utilization During Lookback | | | | |  |  |  |
| Inpatient | | | | | 0.76 | 0.74 | 0.79 |
| Outpatient | | | | | 1.02 | 1.02 | 1.02 |
| Average Number of Diagnoses Per Outpatient Encounter | | | | | 1.19 | 1.17 | 1.20 |
| Elixhauser/AHRQ Comorbidities | | | | |  |  |  |
| Alcohol Abuse | | | | | 0.96 | 0.77 | 1.19 |
| Anemia | | | | | 0.99 | 0.96 | 1.02 |
| Blood Loss Amenia | | | | | 0.84 | 0.79 | 0.90 |
| Coagulopathy | | | | | 0.99 | 0.95 | 1.04 |
| COPD | | | | | 0.83 | 0.81 | 0.85 |
| Depression | | | | | 1.05 | 0.99 | 1.11 |
| Diabetes without Complications | | | | | 0.94 | 0.91 | 0.96 |
| Diabetes with Complications | | | | | 1.04 | 1.00 | 1.08 |
| Drug Abuse | | | | | 1.14 | 1.01 | 1.28 |
| Fluid and Electrolyte Disorders | | | | | 0.98 | 0.96 | 1.02 |
| Heart Failure | | | | | 0.77 | 0.74 | 0.80 |
| HIV | | | | | 0.67 | 0.52 | 0.88 |
| Hypertension without Complications | | | | | 1.08 | 1.05 | 1.10 |
| Hypertension with Complications | | | | | 0.98 | 0.95 | 1.01 |
| Hypothyroidism | | | | | 1.10 | 1.07 | 1.13 |
| Liver Disorders | | | | | 0.93 | 0.88 | 0.97 |
| Lymphoma | | | | | 0.82 | 0.75 | 0.89 |
| Metastatic Cancer | | | | | 0.52 | 0.48 | 0.56 |
| Neurological Disorders and Stroke | | | | | 2.79 | 2.71 | 2.88 |
| Obesity | | | | | 0.93 | 0.90 | 0.96 |
| Paralysis | | | | | 1.31 | 1.24 | 1.39 |
| Peptic Ulcer Disease | | | | | 0.87 | 0.77 | 0.98 |
| Peripheral Vascular Disease | | | | | 1.02 | 1.00 | 1.05 |
| Psychoses | | | | | 1.29 | 1.22 | 1.36 |
| Pulmonary Hypertension | | | | | 0.94 | 0.89 | 0.99 |
| Renal Disorders | | | | | 0.77 | 0.74 | 0.81 |
| Rheumatic Disorders | | | | | 1.14 | 1.10 | 1.19 |
| Solid Tumor | | | | | 0.96 | 0.93 | 0.98 |
| Valvular Disorders | | | | | 1.08 | 1.05 | 1.11 |
| Weight Loss | | | | | 1.34 | 1.28 | 1.39 |
| Clinical Classification Software Codes | | | | |  |  |  |
| Anxiety disorders (651) | | | | | 1.40 | 1.35 | 1.44 |
| Attention-deficit, conduct, and disruptive behavior disorders (652) | | | | | 0.87 | 0.78 | 0.97 |
| Delirium, dementia, and amnestic and other cognitive disorders (653) | | | | | 1.71 | 1.65 | 1.78 |
| Developmental disorders (654) | | | | | 1.26 | 1.10 | 1.46 |
| Disorders usually diagnosed in infancy, childhood, or adolescence (655) | | | | | 1.46 | 1.19 | 1.79 |
| Impulse control disorders, NEC (656) | | | | | 0.65 | 0.48 | 0.88 |
| Mood disorders (657) | | | | | 1.46 | 1.37 | 1.56 |
| Personality disorders (658) | | | | | 0.97 | 0.82 | 1.15 |
| Schizophrenia and other psychotic disorders (659) | | | | | 0.98 | 0.91 | 1.05 |
| Alcohol-related disorders (660) | | | | | 0.89 | 0.72 | 1.10 |
| Substance-related disorders (661) | | | | | 0.73 | 0.67 | 0.79 |
| Suicide and intentional self-inflicted injury (662) | | | | | 1.42 | 1.20 | 1.67 |
| Screening and history of mental health and substance abuse codes (663) | | | | | 0.73 | 0.71 | 0.76 |
| Miscellaneous mental health disorders (670) | | | | | 1.17 | 1.12 | 1.23 |

**Table S6: Full Model Estimate for Ever Use of Sensitivity Medications.** Models were estimated using a logistic fixed effects regression and included a fixed effects for the sampling strata (1 case and 10 controls matched exactly on age, sex, and enrollment period). As a result of the matching, we do not include age, sex, or time as covariates in the model; the fixed effect for the strata addresses confounding by these variables. Standard errors were clustered by the case/control strata.

| **Term** | **Metoclo-pramide** | **Ondanset-ron** | **Bupropion** | **Trazodone** | **Z Drugs** |
| --- | --- | --- | --- | --- | --- |
| Ever Use | 1.33  (1.27 – 1.39) | 0.89  (0.85 – 0.94) | 1.21  (1.16 – 1.27) | 1.05  (1.00 – 1.10) | 1.01  (0.98 – 1.04) |
| Rates of Health Care Utilization During Lookback |  |  |  |  |  |
| Inpatient | 0.77  (0.75 – 0.80) | 0.78  (0.75 – 0.81) | 0.78  (0.75 – 0.81) | 0.78  (0.75 – 0.81) | 0.78  (0.75 – 0.81) |
| Outpatient | 1.02  (1.02 – 1.02) | 1.02  (1.02 – 1.02) | 1.02  (1.02 – 1.02) | 1.02  (1.02 – 1.02) | 1.02  (1.02 – 1.02) |
| Average Number of Diagnoses Per Outpatient Encounter | 1.19  (1.17 – 1.20) | 1.19  (1.17 – 1.20) | 1.19  (1.17 – 1.20) | 1.18  (1.17 – 1.20) | 1.18  (1.17 – 1.20) |
| Elixhauser/AHRQ Comorbidities |  |  |  |  |  |
| Alcohol Abuse | 0.98  (0.78 – 1.22) | 0.97  (0.77 – 1.21) | 0.97  (0.77 – 1.20) | 0.97  (0.77 – 1.20) | 0.97  (0.78 – 1.21) |
| Anemia | 0.99  (0.96 – 1.02) | 0.99  (0.97 – 1.02) | 0.99  (0.97 – 1.02) | 0.99  (0.97 – 1.02) | 0.99  (0.97 – 1.02) |
| Blood Loss Amenia | 0.84  (0.78 – 0.89) | 0.84  (0.78 – 0.90) | 0.84  (0.78 – 0.90) | 0.84  (0.78 – 0.90) | 0.84  (0.78 – 0.90) |
| Coagulopathy | 0.99  (0.95 – 1.04) | 0.99  (0.95 – 1.04) | 0.99  (0.95 – 1.04) | 0.99  (0.95 – 1.04) | 0.99  (0.95 – 1.04) |
| COPD | 0.83  (0.81 – 0.85) | 0.84  (0.82 – 0.86) | 0.83  (0.81 – 0.85) | 0.83  (0.81 – 0.85) | 0.83  (0.81 – 0.85) |
| Depression | 1.04  (0.98 – 1.10) | 1.04  (0.98 – 1.10) | 1.02  (0.96 – 1.08) | 1.04  (0.98 – 1.10) | 1.04  (0.98 – 1.10) |
| Diabetes without Complications | 0.94  (0.92 – 0.97) | 0.94  (0.92 – 0.97) | 0.94  (0.92 – 0.97) | 0.94  (0.92 – 0.97) | 0.94  (0.92 – 0.97) |
| Diabetes with Complications | 1.03  (0.99 – 1.07) | 1.03  (1.00 – 1.07) | 1.03  (1.00 – 1.07) | 1.03  (1.00 – 1.07) | 1.03  (1.00 – 1.07) |
| Drug Abuse | 1.14  (1.01 – 1.28) | 1.15  (1.02 – 1.30) | 1.15  (1.02 – 1.29) | 1.14  (1.01 – 1.28) | 1.14  (1.02 – 1.29) |
| Fluid and Electrolyte Disorders | 0.99  (0.96 – 1.07) | 1.00  (0.97 – 1.03) | 0.99  (0.96 – 1.02) | 0.99  (0.96 – 1.02) | 0.99  (0.96 – 1.02) |
| Heart Failure | 0.77  (0.84 – 0.80) | 0.77  (0.74 – 0.80) | 0.77  (0.74 – 0.80) | 0.77  (0.74 – 0.80) | 0.77  (0.74 – 0.80) |
| HIV | 0.67  (0.52 – 0.88) | 0.68  (0.52 – 0.88) | 0.67  (0.51 – 0.88) | 0.68  (0.52 – 0.88) | 0.68  (0.52 – 0.88) |
| Hypertension without Complications | 1.08  (1.05 – 1.10) | 1.08  (1.05 – 1.10) | 1.08  (1.05 – 1.10) | 1.08  (1.05 – 1.10) | 1.08  (1.05 – 1.10) |
| Hypertension with Complications | 0.98  (0.95 – 1.01) | 0.98  (0.95 – 1.01) | 0.98  (0.95 – 1.01) | 0.98  (0.95 – 1.01) | 0.98  (0.95 – 1.01) |
| Hypothyroidism | 1.10  (1.07 – 1.13) | 1.10  (1.07 – 1.13) | 1.10  (1.07 – 1.13) | 1.10  (1.07 – 1.13) | 1.10  (1.07 – 1.13) |
| Liver Disorders | 0.92  (0.88 – 0.97) | 0.93  (0.89 – 0.98) | 0.93  (0.89 – 0.98) | 0.93  (0.89 – 0.98) | 0.93  (0.89 – 0.98) |
| Lymphoma | 0.85  (0.78 – 0.92) | 0.86  (0.79 – 0.93) | 0.85  (0.79 – 0.92) | 0.85  (0.79 – 0.92) | 0.85  (0.79 – 0.92) |
| Metastatic Cancer | 0.56  (0.52 – 0.60) | 0.57  (0.53 – 0.61) | 0.56  (0.52 – 0.60) | 0.56  (0.52 – 0.60) | 0.56  (0.52 – 0.60) |
| Neurological Disorders and Stroke | 2.82  (2.73 – 2.91) | 2.82  (2.74 – 2.91) | 2.82  (2.74 – 2.91) | 2.82  (2.73 – 2.91) | 2.82  (2.74 – 2.91) |
| Obesity | 0.92  (0.89 – 0.96) | 0.92  (0.89 – 0.96) | 0.92  (0.89 – 0.95) | 0.92  (0.89 – 0.96) | 0.92  (0.89 – 0.96) |
| Paralysis | 1.30  (1.23 – 1.38) | 1.30  (1.23 – 1.38) | 1.30  (1.23 – 1.38) | 1.30  (1.23 – 1.38) | 1.30  (1.23 – 1.38) |
| Peptic Ulcer Disease | 0.86  (0.76 – 0.96) | 0.87  (0.77 – 0.98) | 0.86  (0.77 – 0.97) | 0.86  (0.77 – 0.98) | 0.86  (0.77 – 0.98) |
| Peripheral Vascular Disease | 1.02  (0.99 – 1.05) | 1.02  (1.00 – 1.05) | 1.02  (1.00 – 1.05) | 1.02  (1.00 – 1.05) | 1.02  (1.00 – 1.05) |
| Psychoses | 1.34  (1.27 – 1.42) | 1.35  (1.27 – 1.42) | 1.31  (1.24 – 1.39) | 1.34  (1.27 – 1.42) | 1.34  (1.27 – 1.42) |
| Pulmonary Hypertension | 0.94  (0.89 – 0.99) | 0.94  (0.89 – 0.99) | 0.94  (0.89 – 0.99) | 0.94  (0.89 – 0.99) | 0.94  (0.89 – 0.99) |
| Renal Disorders | 0.77  (0.74 – 0.80) | 0.77  (0.74 – 0.80) | 0.77  (0.74 – 0.80) | 0.77  (0.74 – 0.80) | 0.77  (0.74 – 0.80) |
| Rheumatic Disorders | 1.14  (1.10 – 1.18) | 1.14  (1.10 – 1.18) | 1.14  (1.10 – 1.18) | 1.14  (1.10 – 1.18) | 1.14  (1.10 – 1.18) |
| Solid Tumor | 0.96  (0.94 – 0.99) | 0.97  (0.94 – 0.99) | 0.96  (0.94 – 0.99) | 0.96  (0.94 – 0.99) | 0.96  (0.94 – 0.99) |
| Valvular Disorders | 1.08  (1.05 – 1.11) | 1.08  (1.05 – 1.11) | 1.08  (1.05 – 1.11) | 1.08  (1.05 – 1.11) | 1.08  (1.05 – 1.11) |
| Weight Loss | 1.33  (1.28 – 1.39) | 1.35  (1.30 – 1.40) | 1.34  (1.29 – 1.40) | 1.34  (1.29 – 1.40) | 1.34  (1.29 – 1.40) |
| Clinical Classification Software Codes |  |  |  |  |  |
| Anxiety disorders (651) | 1.41  (1.37 – 1.46) | 1.42  (1.37 – 1.46) | 1.41  (1.36 – 1.45) | 1.41  (1.37 – 1.46) | 1.41  (1.37 – 1.46) |
| Attention-deficit, conduct, and disruptive behavior disorders (652) | 0.90  (0.81 – 1.00) | 0.89  (0.80 – 0.99) | 0.88  (0.79 – 0.98) | 0.89  (0.80 – 0.99) | 0.89  (0.80 – 0.99) |
| Delirium, dementia, and amnestic and other cognitive disorders (653) | 1.81  (1.74 – 1.88) | 1.80  (1.73 – 1.87) | 1.80  (1.74 – 1.87) | 1.80  (1.73 – 1.87) | 1.80  (1.74 – 1.87) |
| Developmental disorders (654) | 1.33  (1.16 – 1.53) | 1.33  (1.15 – 1.52) | 1.33  (1.16 – 1.53) | 1.32  (1.15 – 1.52) | 1.33  (1.15 – 1.53) |
| Disorders usually diagnosed in infancy, childhood, or adolescence (655) | 1.47  (1.20 – 1.81) | 1.47  (1.20 – 1.81) | 1.46  (1.18 – 1.80) | 1.47  (1.19 – 1.81) | 1.47  (1.19 – 1.81) |
| Impulse control disorders, NEC (656) | 0.68  (0.50 – 0.93) | 0.68  (0.50 – 0.92) | 0.68  (0.50 – 0.93) | 0.68  (0.50 – 0.92) | 0.68  (0.50 – 0.93) |
| Mood disorders (657) | 1.50  (1.41 – 1.60) | 1.50  (1.41 – 1.60) | 1.49  (1.40 – 1.59) | 1.50  (1.41 – 1.60) | 1.50  (1.41 – 1.60) |
| Personality disorders (658) | 1.03  (0.87 – 1.22) | 1.02  (0.86 – 1.21) | 1.02  (0.86 – 1.21) | 1.02  (0.86 – 1.21) | 1.02  (0.86 – 1.21) |
| Schizophrenia and other psychotic disorders (659) | 1.02  (0.95 – 1.09) | 1.02  (0.95 – 1.09) | 1.04  (0.97 – 1.12) | 1.02  (0.95 – 1.09) | 1.02  (0.95 – 1.09) |
| Alcohol-related disorders (660) | 0.88  (0.71 – 1.08) | 0.88  (0.71 – 1.08) | 0.88  (0.71 – 1.08) | 0.88  (0.71 – 1.08) | 0.88  (0.71 – 1.08) |
| Substance-related disorders (661) | 0.73  (0.68 – 0.79) | 0.73  (0.68 – 0.80) | 0.73  (0.67 – 0.79) | 0.73  (0.68 – 0.79) | 0.73  (0.68 – 0.79) |
| Suicide and intentional self-inflicted injury (662) | 1.50  (1.27 – 1.76) | 1.49  (1.27 – 1.76) | 1.46  (1.24 – 1.73) | 1.48  (1.26 – 1.74) | 1.49  (1.26 – 1.75) |
| Screening and history of mental health and substance abuse codes (663) | 0.73  (0.71 – 0.76) | 0.73  (0.71 – 0.76) | 0.73  (0.70 – 0.76) | 0.73  (0.71 – 0.76) | 0.73  (0.71 – 0.76) |
| Miscellaneous mental health disorders (670) | 1.17  (1.12 – 1.23) | 1.18  (1.13 – 1.23) | 1.17  (1.12 – 1.23) | 1.17  (1.12 – 1.23) | 1.18  (1.12 – 1.23) |

**Table S7: Full Model Estimate for Days Supplied of Sensitivity Medications.** Models were estimated using a logistic fixed effects regression and included a fixed effects for the sampling strata (1 case and 10 controls matched exactly on age, sex, and enrollment period). As a result of the matching, we do not include age, sex, or time as covariates in the model; the fixed effect for the strata addresses confounding by these variables. Standard errors were clustered by the case/control strata.

| **Term** | **Metoclo-pramide** | **Ondanset-ron** | **Bupropion** | **Trazodone** | **Z Drugs** |
| --- | --- | --- | --- | --- | --- |
| Days Supplied |  |  |  |  |  |
| Never User | Reference | Reference | Reference | Reference | Reference |
| < 1 Month | 0.98  (0.91 – 1.05) | 0.90  (0.85 – 0.94) | 1.45  (1.16 – 1.82) | 1.11  (0.94 – 1.32) | 1.06  (0.98 – 1.14) |
| >= 1 Month and < 3 Months | 1.16  (1.06 – 1.26) | 0.83  (0.73 – 0.96) | 1.20  (1.10 – 1.30) | 1.07  (0.99 – 1.15) | 1.09  (1.04 – 1.15) |
| >= 3 Months and < 6 Months | 1.63  (1.42 – 1.86) | 0.83  (0.62 – 1.11) | 1.20  (1.08 – 1.34) | 1.09  (0.97 – 1.21) | 1.09  (1.01 – 1.17) |
| >= 6 Months and < 9 Months | 2.02  (1.66 – 2.45) | 1.14  (0.67 – 1.94) | 1.33  (1.15 – 1.53) | 0.97  (0.84 – 1.13) | 1.02  (0.92 – 1.12) |
| >= 9 Months and < 1 Year | 2.64  (2.15 – 3.26) | 1.02  (0.54 – 1.96) | 1.26  (1.09 – 1.46) | 1.20  (1.03 – 1.39) | 0.94  (0.84 – 1.05) |
| >= 1 Year and < 2 Years | 3.01  (2.54 – 3.56) | 0.96  (0.50 – 1.82) | 1.32  (1.19 – 1.47) | 1.04  (0.93 – 1.17) | 0.95  (0.88 – 1.03) |
| >= 2 Years and < 3 Years | 2.77  (2.17 – 3.55) | 1.77  (0.45 – 6.89) | 1.11  (0.96 – 1.29) | 1.00  (0.85 – 1.18) | 0.83  (0.74 – 0.93) |
| >= 3 Years | 2.71  (2.14 – 3.42) | 2.37  (0.39 – 14.25) | 1.08  (0.97 – 1.20) | 0.92  (0.81 – 1.05) | 0.85  (0.78 – 0.93) |
| Rates of Health Care Utilization During Lookback |  |  |  |  |  |
| Inpatient | 0.77  (0.74 – 0.80) | 0.78  (0.75 – 0.81) | 0.78  (0.75 – 0.81) | 0.78  (0.75 – 0.81) | 0.78  (0.75 – 0.81) |
| Outpatient | 1.02  (1.02 – 1.02) | 1.02  (1.02 – 1.02) | 1.02  (1.02 – 1.02) | 1.02  (1.02 – 1.02) | 1.02  (1.02 – 1.02) |
| Average Number of Diagnoses Per Outpatient Encounter | 1.19  (1.17 – 1.20) | 1.19  (1.17 – 1.20) | 1.19  (1.17 – 1.20) | 1.18  (1.17 – 1.20) | 1.18  (1.17 – 1.20) |
| Elixhauser/AHRQ Comorbidities |  |  |  |  |  |
| Alcohol Abuse | 0.98  (0.78 – 1.22) | 0.97  (0.78 – 1.21) | 0.97  (0.77 – 1.20) | 0.97  (0.77 – 1.21) | 0.97  (0.77 – 1.20) |
| Anemia | 0.99  (0.96 – 1.02) | 0.99  (0.97 – 1.02) | 0.99  (0.97 – 1.02) | 0.99  (0.97 – 1.02) | 0.99  (0.97 – 1.02) |
| Blood Loss Amenia | 0.83  (0.78 – 0.89) | 0.84  (0.78 – 0.90) | 0.84  (0.78 – 0.90) | 0.84  (0.78 – 0.90) | 0.84  (0.79 – 0.90) |
| Coagulopathy | 0.99  (0.85 – 1.04) | 0.99  (0.95 – 1.04) | 0.99  (0.95 – 1.04) | 0.99  (0.95 – 1.04) | 0.99  (0.85 – 1.04) |
| COPD | 0.83  (0.81 – 0.85) | 0.84  (0.82 – 0.86) | 0.83  (0.81 – 0.85) | 0.83  (0.81 – 0.85) | 0.83  (0.81 – 0.85) |
| Depression | 1.04  (0.98 – 1.10) | 1.04  (0.98 – 1.10) | 1.02  (0.96 – 1.09) | 1.04  (0.98 – 1.10) | 1.04  (0.98 – 1.10) |
| Diabetes without Complications | 0.94  (0.91 – 0.96) | 0.94  (0.92 – 0.97) | 0.94  (0.92 – 0.97) | 0.94  (0.92 – 0.97) | 0.94  (0.92 – 0.97) |
| Diabetes with Complications | 1.02  (0.99 – 1.06) | 1.03  (1.00 – 1.07) | 1.03  (1.00 – 1.07) | 1.03  (1.00 – 1.07) | 1.03  (1.00 – 1.07) |
| Drug Abuse | 1.14  (1.01 – 1.28) | 1.15  (1.02 – 1.30) | 1.15  (1.02 – 1.29) | 1.14  (1.01 – 1.28) | 1.15  (1.03 – 1.30) |
| Fluid and Electrolyte Disorders | 0.99  (0.96 – 1.02) | 1.00  (0.97 – 1.03) | 0.99  (0.96 – 1.02) | 0.99  (0.96 – 1.02) | 0.99  (0.96 – 1.02) |
| Heart Failure | 0.77  (0.74 – 0.79) | 0.77  (0.74 – 0.80) | 0.77  (0.74 – 0.80) | 0.77  (0.74 – 0.80) | 0.77  (0.74 – 0.80) |
| HIV | 0.67  (0.52 – 0.88) | 0.68  (0.52 – 0.89) | 0.67  (0.51 – 0.88) | 0.68  (0.52 – 0.88) | 0.68  (0.52 – 0.89) |
| Hypertension without Complications | 1.08  (1.05 – 1.10) | 1.08  (1.05 – 1.10) | 1.08  (1.05 – 1.10) | 1.08  (1.05 – 1.10) | 1.08  (1.05 – 1.10) |
| Hypertension with Complications | 0.98  (0.95 – 1.01) | 0.98  (0.95 – 1.01) | 0.98  (0.95 – 1.01) | 0.98  (0.95 – 1.01) | 0.98  (0.95 – 1.01) |
| Hypothyroidism | 1.10  (1.07 – 1.13) | 1.10  (1.07 – 1.13) | 1.10  (1.07 – 1.13) | 1.10  (1.07 – 1.13) | 1.10  (1.07 – 1.13) |
| Liver Disorders | 0.92  (0.88 – 0.97) | 0.93  (0.89 – 0.98) | 0.93  (0.89 – 0.98) | 0.93  (0.89 – 0.98) | 0.93  (0.89 – 0.98) |
| Lymphoma | 0.86  (0.79 – 0.93) | 0.86  (0.79 – 0.93) | 0.85  (0.79 – 0.92) | 0.85  (0.79 – 0.92) | 0.85  (0.78 – 0.92) |
| Metastatic Cancer | 0.56  (0.52 – 0.60) | 0.57  (0.53 – 0.61) | 0.56  (0.52 – 0.61) | 0.56  (0.52 – 0.60) | 0.56  (0.52 – 0.60) |
| Neurological Disorders and Stroke | 2.82  (2.73 – 2.91) | 2.82  (2.74 – 2.91) | 2.82  (2.74 – 2.91) | 2.82  (2.72 – 2.91) | 2.82  (2.74 – 2.91) |
| Obesity | 0.93  (0.89 – 0.96) | 0.92  (0.89 – 0.96) | 0.92  (0.89 – 0.95) | 0.92  (0.89 – 0.95) | 0.92  (0.89 – 0.96) |
| Paralysis | 1.30  (1.23 – 1.38) | 1.30  (1.23 – 1.38) | 1.30  (1.23 – 1.38) | 1.30  (1.23 – 1.38) | 1.30  (1.23 – 1.38) |
| Peptic Ulcer Disease | 0.85  (0.76 – 0.96) | 0.87  (0.77 – 0.98) | 0.86  (0.77 – 0.97) | 0.86  (0.77 – 0.97) | 0.87  (0.77 – 0.98) |
| Peripheral Vascular Disease | 1.02  (0.99 – 1.05) | 1.02  (1.00 – 1.05) | 1.02  (1.00 – 1.05) | 1.02  (1.00 – 1.05) | 1.02  (1.00 – 1.05) |
| Psychoses | 1.34  (1.27 – 1.42) | 1.35  (1.27 – 1.42) | 1.32  (1.24 – 1.39) | 1.34  (1.27 – 1.42) | 1.35  (1.28 – 1.42) |
| Pulmonary Hypertension | 0.93  (0.88 – 0.98) | 0.94  (0.89 – 0.99) | 0.94  (0.89 – 0.99) | 0.94  (0.89 – 0.99) | 0.94  (0.89 – 0.99) |
| Renal Disorders | 0.77  (0.74 – 0.80) | 0.77  (0.74 – 0.80) | 0.77  (0.74 – 0.80) | 0.77  (0.74 – 0.80) | 0.77  (0.74 – 0.80) |
| Rheumatic Disorders | 1.13  (1.09 – 1.18) | 1.14  (1.10 – 1.18) | 1.14  (1.10 – 1.18) | 1.14  (1.10 – 1.18) | 1.14  (1.10 – 1.18) |
| Solid Tumor | 0.96  (0.94 – 0.99) | 0.97  (0.94 – 0.99) | 0.96  (0.94 – 0.99) | 0.96  (0.94 – 0.99) | 0.96  (0.94 – 0.99) |
| Valvular Disorders | 1.08  (1.05 – 1.11) | 1.08  (1.05 – 1.11) | 1.08  (1.05 – 1.11) | 1.08  (1.05 – 1.11) | 1.08  (1.05 – 1.11) |
| Weight Loss | 1.33  (1.27 – 1.38) | 1.35  (1.30 – 1.40) | 1.34  (1.29 – 1.40) | 1.34  (1.29 – 1.40) | 1.35  (1.29 – 1.40) |
| Clinical Classification Software Codes |  |  |  |  |  |
| Anxiety disorders (651) | 1.41  (1.37 – 1.45) | 1.42  (1.37 – 1.46) | 1.41  (1.36 – 1.45) | 1.41  (1.37 – 1.46) | 1.42  (1.37 – 1.46) |
| Attention-deficit, conduct, and disruptive behavior disorders (652) | 0.90  (0.81 – 1.00) | 0.89  (0.80 – 0.99) | 0.88  (0.79 – 0.98) | 0.89  (0.80 – 0.99) | 0.89  (0.80 – 0.99) |
| Delirium, dementia, and amnestic and other cognitive disorders (653) | 1.81  (1.74 – 1.88) | 1.80  (1.73 – 1.87) | 1.80  (1.74 – 1.87) | 1.80  (1.73 – 1.87) | 1.80  (1.73 – 1.87) |
| Developmental disorders (654) | 1.33  (1.16 – 1.53) | 1.32  (1.15 – 1.52) | 1.33  (1.16 – 1.53) | 1.32  (1.15 – 1.52) | 1.32  (1.15 – 1.52) |
| Disorders usually diagnosed in infancy, childhood, or adolescence (655) | 1.47  (1.20 – 1.81) | 1.47  (1.20 – 1.81) | 1.46  (1.18 – 1.80) | 1.47  (1.19 – 1.81) | 1.47  (1.19 – 1.81) |
| Impulse control disorders, NEC (656) | 0.68  (0.50 – 0.92) | 0.68  (0.49 – 0.92) | 0.68  (0.50 – 0.93) | 0.68  (0.50 – 0.92) | 0.68  (0.50 – 0.92) |
| Mood disorders (657) | 1.50  (1.40 – 1.60) | 1.50  (1.41 – 1.60) | 1.49  (1.40 – 1.59) | 1.50  (1.41 – 1.60) | 1.50  (1.41 – 1.60) |
| Personality disorders (658) | 1.03  (0.87 – 1.22) | 1.02  (0.86 – 1.21) | 1.02  (0.86 – 1.21) | 1.02  (0.86 – 1.21) | 1.02  (0.86 – 1.21) |
| Schizophrenia and other psychotic disorders (659) | 1.02  (0.95 – 1.10) | 1.02  (0.95 – 1.09) | 1.04  (0.97 – 1.11) | 1.02  (0.95 – 1.09) | 1.02  (0.95 – 1.09) |
| Alcohol-related disorders (660) | 0.88  (0.71 – 1.09) | 0.88  (0.71 – 1.08) | 0.88  (0.71 – 1.08) | 0.88  (0.71 – 1.08) | 0.88  (0.71 – 1.09) |
| Substance-related disorders (661) | 0.73  (0.68 – 0.79) | 0.74  (0.68 – 0.80) | 0.73  (0.67 – 0.79) | 0.73  (0.68 – 0.79) | 0.73  (0.68 – 0.79) |
| Suicide and intentional self-inflicted injury (662) | 1.50  (1.27 – 1.76) | 1.49  (1.27 – 1.76) | 1.46  (1.24 – 1.73) | 1.48  (1.26 – 1.74) | 1.49  (1.26 – 1.76) |
| Screening and history of mental health and substance abuse codes (663) | 0.74  (0.71 – 0.76) | 0.73  (0.71 – 0.76) | 0.73  (0.70 – 0.76) | 0.73  (0.71 – 0.76) | 0.73  (0.71 – 0.76) |
| Miscellaneous mental health disorders (670) | 1.17  (1.12 – 1.23) | 1.18  (1.13 – 1.23) | 1.17  (1.12 – 1.23) | 1.17  (1.12 – 1.23) | 1.18  (1.13 – 1.24) |

**Table S8: Models to Address Potential Collider Bias and Confounder by Indication.** In the model removing potential colliders, the collider variables were omitted at the time of estimation and the full model data set was used. In the time windowing analysis, the potential collider comorbidities were calculated using data excluding the period 1, 3, or 5 years prior to the PD index date. To ensure these comorbidities could be calculated, we also excluded any strata without at one day of lookback prior to the time windowing threshold and so the sample size varies as reported in the table.

| **Term** | **Remove Potential Colliders (N = 717,639)** | **Time Windowing 1 Year (N = 685,158)** | **Time Windowing 3 Years (N = 406,082)** | **Time Windowing 5 Years (N = 254,275)** |
| --- | --- | --- | --- | --- |
| Ever Use | 1.81  (1.74 – 1.87) | 1.60  (1.54 – 1.67) | 1.52  (1.44 – 1.61) | 1.55  (1.44 – 1.67) |
| Rates of Health Care Utilization During Lookback |  |  |  |  |
| Inpatient | 0.88  (0.85 – 0.91) | 0.88  (0.85 – 0.91) | 0.89  (0.84 – 0.95) | 0.89  (0.82 – 0.98) |
| Outpatient | 1.02  (1.02 – 1.02) | 1.02  (1.02 – 1.02) | 1.03  (1.02 – 1.03) | 1.03  (1.03 – 1.03) |
| Average Number of Diagnoses Per Outpatient Encounter | 1.24  (1.22 – 1.25) | 1.24  (1.23 – 1.26) | 1.29  (1.26 – 1.31) | 1.27  (1.23 – 1.30) |
| Elixhauser/AHRQ Comorbidities |  |  |  |  |
| Alcohol Abuse | 1.02  (0.82 – 1.27) | 1.06  (0.85 – 1.33) | 1.09  (0.84 – 1.41) | 1.13  (0.83 – 1.55) |
| Anemia | 1.01  (0.99 – 1.04) | 1.01  (0.98 – 1.04) | 1.02  (0.98 – 1.05) | 1.01  (0.98 – 1.06) |
| Blood Loss Amenia | 0.83  (0.78 – 0.89) | 0.83  (0.78 – 0.89) | 0.85  (0.79 – 0.91) | 0.85  (0.77 – 0.93) |
| Coagulopathy | 1.00  (0.96 – 1.05) | 1.00  (0.95 – 1.05) | 1.01  (0.96 – 1.07) | 1.01  (0.95 – 1.08) |
| COPD | 0.83  (0.81 – 0.85) | 0.84  (0.82 – 0.86) | 0.85  (0.83 – 0.88) | 0.86  (0.83 – 0.89) |
| Depression | --- | 0.95  (0.89 – 1.02) | 1.00  (0.91 – 1.09) | 0.93  (0.82 – 1.05) |
| Diabetes without Complications | 0.93  (0.91 – 0.96) | 0.93  (0.91 – 0.96) | 0.93  (0.90 – 0.96) | 0.93  (0.89 – 0.96) |
| Diabetes with Complications | 1.06  (1.03 – 1.10) | 1.04  (1.01 – 1.08) | 1.03  (0.99 – 1.08) | 1.02  (0.97 – 1.08) |
| Drug Abuse | 1.09  (0.97 – 1.23) | 1.14  (1.01 – 1.29) | 1.08  (0.94 – 1.25) | 1.02  (0.86 – 1.21) |
| Fluid and Electrolyte Disorders | 1.08  (1.04 – 1.11) | 1.06  (1.03 – 1.09) | 1.09  (1.05 – 1.13) | 1.09  (1.04 – 1.14) |
| Heart Failure | 0.78  (0.76 – 0.81) | 0.78  (0.75 – 0.81) | 0.79  (0.76 – 0.83) | 0.80  (0.76 – 0.84) |
| HIV | 0.68  (0.52 – 0.88) | 0.67  (0.51 – 0.88) | 0.70  (0.50 – 0.96) | 0.69  (0.47 – 1.04) |
| Hypertension without Complications | 1.07  (1.05 – 1.10) | 1.08  (1.05 – 1.10) | 1.06  (1.03 – 1.09) | 1.06  (1.02 – 1.10) |
| Hypertension with Complications | 0.99  (0.96 – 1.02) | 0.97  (0.94 – 1.00) | 0.97  (0.94 – 1.01) | 0.99  (0.94 – 1.03) |
| Hypothyroidism | 1.11  (1.08 – 1.14) | 1.11  (1.08 – 1.14) | 1.11  (1.07 – 1.14) | 1.10  (1.06 – 1.14) |
| Liver Disorders | 0.92  (0.88 – 0.96) | 0.92  (0.88 – 0.97) | 0.94  (0.89 – 0.99) | 0.98  (0.91 – 1.04) |
| Lymphoma | 0.78  (0.72 – 0.84) | 0.82  (0.75 – 0.89) | 0.85  (0.77 – 0.94) | 0.89  (0.79 – 1.00) |
| Metastatic Cancer | 0.49  (0.46 – 0.53) | 0.55  (0.51 – 0.60) | 0.61  (0.56 – 0.66) | 0.62  (0.56 – 0.69) |
| Neurological Disorders and Stroke | --- | 1.82  (1.75 – 1.88) | 1.45  (1.37 – 1.52) | 1.35  (1.25 – 1.45) |
| Obesity | 0.90  (0.87 – 0.93) | 0.92  (0.89 – 0.95) | 0.93  (0.90 – 0.97) | 0.93  (0.89 – 0.97) |
| Paralysis | --- | 0.86  (0.80 – 0.93) | 0.71  (0.63 – 0.79) | 0.71  (0.60 – 0.83) |
| Peptic Ulcer Disease | 0.87  (0.77 – 0.97) | 0.88  (0.78 – 0.99) | 0.90  (0.78 – 1.03) | 0.94  (0.81 – 1.10) |
| Peripheral Vascular Disease | 1.07  (1.04 – 1.10) | 1.05  (1.02 – 1.08) | 1.06  (1.03 – 1.10) | 1.07  (1.03 – 1.11) |
| Psychoses | --- | 1.24  (1.17 – 1.33) | 1.24  (1.13 – 1.36) | 1.17  (1.03 – 1.33) |
| Pulmonary Hypertension | 0.94  (0.90 – 0.99) | 0.93  (0.88 – 0.98) | 0.93  (0.87 – 0.98) | 0.93  (0.87 – 1.00) |
| Renal Disorders | 0.78  (0.75 – 0.81) | 0.77  (0.74 – 0.80) | 0.78  (0.75 – 0.82) | 0.79  (0.75 – 0.84) |
| Rheumatic Disorders | 1.13  (1.09 – 1.17) | 1.14  (1.10 – 1.18) | 1.13  (1.08 – 1.18) | 1.14  (1.09 – 1.20) |
| Solid Tumor | 0.94  (0.91 – 0.96) | 0.95  (0.93 – 0.98) | 0.98  (0.95 – 1.01) | 0.97  (0.93 – 1.01) |
| Valvular Disorders | 1.10  (1.07 – 1.13) | 1.09  (1.06 – 1.12) | 1.10  (1.07 – 1.14) | 1.09  (1.05 – 1.13) |
| Weight Loss | 1.46  (1.41 – 1.52) | 1.49  (1.43 – 1.55) | 1.54  (1.48 – 1.62) | 1.53  (1.45 – 1.62) |
| Clinical Classification Software Codes |  |  |  |  |
| Anxiety disorders (651) | 1.43  (1.39 – 1.47) | 1.21  (1.16 – 1.25) | 1.13  (1.07 – 1.18) | 1.09  (1.02 – 1.17) |
| Attention-deficit, conduct, and disruptive behavior disorders (652) | 0.97  (0.87 – 1.07) | 1.00  (0.90 – 1.12) | 1.16  (1.02 – 1.33) | 1.26  (1.03 – 1.47) |
| Delirium, dementia, and amnestic and other cognitive disorders (653) | --- | 1.38  (1.32 – 1.45) | 1.16  (1.07 – 1.24) | 0.98  (0.88 – 1.11) |
| Developmental disorders (654) |  | 1.79  (1.54 – 2.08) | 1.98  (1.66 – 2.38) | 2.10  (1.69 – 2.60) |
| Disorders usually diagnosed in infancy, childhood, or adolescence (655) |  | 1.53  (1.24 – 1.88) | 1.44  (1.13 – 1.84) | 1.35  (1.01 – 1.81) |
| Impulse control disorders, NEC (656) |  | 0.56  (0.37 – 0.83) | 0.63  (0.37 – 1.08) | 0.78  (0.40 – 1.54) |
| Mood disorders (657) |  | 1.44  (1.33 – 1.55) | 1.25  (1.13 – 1.39) | 1.30  (1.13 – 1.49) |
| Personality disorders (658) |  | 1.08  (0.90 – 1.30) | 1.15  (0.92 – 1.44) | 1.28  (0.98 – 1.67) |
| Schizophrenia and other psychotic disorders (659) |  | 0.97  (0.89 – 1.05) | 1.10  (0.98 – 1.25) | 1.30  (1.09 – 1.54) |
| Alcohol-related disorders (660) |  | 0.88  (0.72 – 1.09) | 0.90  (0.71 – 1.16) | 0.86  (0.64 – 1.17) |
| Substance-related disorders (661) |  | 0.77  (0.71 – 0.83) | 0.81  (0.74 – 0.89) | 0.86  (0.77 – 0.96) |
| Suicide and intentional self-inflicted injury (662) |  | 1.72  (1.44 – 2.04) | 2.00  (1.62 – 2.46) | 2.23  (1.75 – 2.85) |
| Screening and history of mental health and substance abuse codes (663) |  | 0.74  (0.72 – 0.77) | 0.75  (0.72 – 0.78) | 0.75  (0.71 – 0.78) |
| Miscellaneous mental health disorders (670) |  | 1.31  (1.25 – 1.37) | 1.38  (1.31 – 1.45) | 1.32  (1.24 – 1.40) |

**Table S9: Average Marginal Odds Ratio for Ever Use Interacted with Potential Indications for Use of Antipsychotics.** The average marginal odds ratio was calculated to combine the joint main and interactive effects in the models with interactions. The 95% CI were calculated with fractional weighted bootstrapping with 9,999 replicates. Bootstrap weights were drawn by the case/control strata such that all members of a given stratum had the same weights. Weights were drawn from an Exp(1) distribution scaled to have a mean of one. Other than the interactions, the model was adjusted as in the primary analysis for patient factors.

|  | Has Diagnosis | | | Does Not Have Diagnosis | | |
| --- | --- | --- | --- | --- | --- | --- |
|  | Odds Ratio | 95% CI | | Odds Ratio | 95% CI | |
|  |  | Lower Bound | Upper Bound |  | Lower Bound | Upper Bound |
| Any | 1.41 | 1.36 | 1.48 | 1.53 | 1.45 | 1.61 |
| Elixhauser Psychoses | 1.95 | 1.83 | 2.09 | 1.47 | 1.40 | 1.55 |
| Elixhauser Depression | 1.55 | 1.44 | 1.66 | 1.50 | 1.42 | 1.58 |
| Anxiety Disorders (CCS 651) | 1.38 | 1.28 | 1.48 | 1.52 | 1.44 | 1.60 |
| CCS Delirium, Dementia (CCS 653) | 1.00 | 0.93 | 1.07 | 1.55 | 1.47 | 1.63 |
| Impulse Control (CCS 656) | 2.92 | 1.56 | 5.69 | 1.50 | 1.43 | 1.57 |
| Mood Disorders (CCS 657) | 1.73 | 1.63 | 1.84 | 1.47 | 1.40 | 1.55 |
| Personality Disorders (CCS 658) | 2.34 | 1.65 | 3.37 | 1.50 | 1.43 | 1.57 |
| Schizophrenia (CCS 659) | 1.49 | 1.34 | 1.65 | 1.50 | 1.43 | 1.58 |

**Table S10: Estimated Associations Between Ever Use and Development of PD for Medications with 1,000 or More Dispensing Claims.** Adjusted odds ratios are adjusted for matching (age, sex, time), health care utilization during lookback, and comorbid health conditions at the index date. Values in parenthesis and estimated 95% CIs using standard errors clustered at the level of the sampling group (1 case and 10 matched controls).

|  | Unadjusted | | Regression Adjusted | |
| --- | --- | --- | --- | --- |
| Medication | Odds Ratio | 95% CI | Odds Ratio | 95% CI |
| Aripiprazole | 6.26 | 5.89 – 6.65 | 3.45 | 3.13 – 3.80 |
| Chlorpromazine | 1.75 | 1.50 – 2.05 | 1.19 | 0.97 – 1.46 |
| Haloperidol | 3.43 | 3.11 – 3.79 | 1.32 | 1.14 – 1.52 |
| Olanzapine | 4.10 | 3.88 – 4.33 | 1.95 | 1.80 – 2.12 |
| Prochlorperazine | 1.12 | 1.07 – 1.18 | 0.90 | 0.85 – 0.96 |
| Quetiapine | 3.02 | 2.89 – 3.16 | 1.09 | 1.02 – 1.17 |
| Risperidone | 3.80 | 3.62 – 3.99 | 1.61 | 1.49 – 1.73 |
| Ziprasidone | 5.73 | 5.04 – 6.51 | 2.51 | 2.07 – 3.04 |

**Table S11: Estimated Associations Between Days Supplied and Development of PD for Common Medications.** Adjusted odds ratios are adjusted for matching (age, sex, time), health care utilization during lookback, and comorbid health conditions at the index date. Values in parenthesis and estimated 95% CIs using standard errors clustered at the level of the sampling group (1 case and 10 matched controls).

|  |  |  | **95% CI** | |
| --- | --- | --- | --- | --- |
| **Term** | **N Exposed** | **Odds Ratio** | **Lower Bound** | **Upper Bound** |
| Aripiprazole |  |  |  |  |
| Duration of Exposure |  |  |  |  |
| Never User | 713,222 | Reference | | |
| < 1 Month | 182 | 2.26 | 1.39 | 3.68 |
| >= 1 Month and < 3 Months | 1,082 | 2.18 | 1.80 | 2.64 |
| >= 3 Months and < 6 Months | 767 | 2.51 | 2.01 | 3.13 |
| >= 6 Months and < 9 Months | 453 | 3.13 | 2.35 | 4.16 |
| >= 9 Months and < 1 Year | 451 | 5.24 | 3.89 | 7.06 |
| >= 1 Year and < 2 Years | 758 | 5.68 | 4.50 | 7.17 |
| >= 2 Years and < 3 Years | 322 | 5.47 | 3.89 | 7.70 |
| >= 3 Years | 402 | 4.72 | 3.46 | 6.45 |
| Chlorpromazine |  |  |  |  |
| Duration of Exposure |  |  |  |  |
| Never User | 716,365 | Reference | | |
| < 1 Month | 857 | 1.02 | 0.78 | 1.32 |
| >= 1 Month and < 3 Months | 204 | 0.95 | 0.57 | 1.57 |
| >= 3 Months and < 6 Months | 55 | 1.39 | 0.57 | 3.41 |
| >= 6 Months and < 9 Months | 22 | 5.22 | 1.37 | 19.85 |
| >= 9 Months and < 1 Year | 19 | 3.32 | 0.66 | 16.82 |
| >= 1 Year and < 2 Years | 47 | 1.60 | 0.61 | 4.18 |
| >= 2 Years and < 3 Years | 22 | 5.65 | 1.63 | 19.59 |
| >= 3 Years | 48 | 1.58 | 0.56 | 4.45 |
| Haloperidol |  |  |  |  |
| Duration of Exposure |  |  |  |  |
| Never User | 715,526 | Reference | | |
| < 1 Month | 557 | 0.64 | 0.47 | 0.86 |
| >= 1 Month and < 3 Months | 693 | 1.12 | 0.87 | 1.43 |
| >= 3 Months and < 6 Months | 302 | 1.53 | 1.07 | 2.19 |
| >= 6 Months and < 9 Months | 110 | 2.93 | 1.61 | 5.33 |
| >= 9 Months and < 1 Year | 102 | 2.34 | 1.26 | 4.34 |
| >= 1 Year and < 2 Years | 176 | 2.51 | 1.56 | 4.04 |
| >= 2 Years and < 3 Years | 71 | 2.61 | 1.18 | 5.77 |
| >= 3 Years | 102 | 3.59 | 1.92 | 6.70 |
| Olanzapine |  |  |  |  |
| Duration of Exposure |  |  |  |  |
| Never User | 711,041 | Reference | | |
| < 1 Month | 368 | 1.41 | 1.00 | 1.98 |
| >= 1 Month and < 3 Months | 1,677 | 1.41 | 1.20 | 1.65 |
| >= 3 Months and < 6 Months | 1,014 | 1.69 | 1.38 | 2.08 |
| >= 6 Months and < 9 Months | 613 | 2.17 | 1.68 | 2.79 |
| >= 9 Months and < 1 Year | 568 | 2.38 | 1.81 | 3.11 |
| >= 1 Year and < 2 Years | 1,119 | 2.35 | 1.95 | 2.85 |
| >= 2 Years and < 3 Years | 495 | 2.51 | 1.88 | 3.36 |
| >= 3 Years | 744 | 2.81 | 2.24 | 3.53 |
| Prochlorperazine |  |  |  |  |
| Duration of Exposure |  |  |  |  |
| Never User | 699,773 | Reference | | |
| < 1 Month | 15,240 | 0.83 | 0.77 | 0.89 |
| >= 1 Month and < 3 Months | 1,907 | 0.93 | 0.78 | 1.12 |
| >= 3 Months and < 6 Months | 370 | 2.04 | 1.44 | 2.89 |
| >= 6 Months and < 9 Months | 123 | 2.57 | 1.43 | 4.64 |
| >= 9 Months and < 1 Year | 75 | 3.13 | 1.41 | 6.95 |
| >= 1 Year and < 2 Years | 94 | 2.14 | 1.06 | 4.29 |
| >= 2 Years and < 3 Years | 31 | 26.98 | 7.55 | 96.42 |
| >= 3 Years | 26 | 1.83 | 0.46 | 7.20 |
| Quetiapine |  |  |  |  |
| Duration of Exposure |  |  |  |  |
| Never User | 706,984 | Reference | | |
| < 1 Month | 653 | 1.22 | 0.95 | 1.57 |
| >= 1 Month and < 3 Months | 2,930 | 1.39 | 1.23 | 1.57 |
| >= 3 Months and < 6 Months | 1,568 | 1.16 | 0.99 | 1.37 |
| >= 6 Months and < 9 Months | 896 | 1.11 | 0.89 | 1.39 |
| >= 9 Months and < 1 Year | 871 | 1.09 | 0.87 | 1.36 |
| >= 1 Year and < 2 Years | 1,692 | 0.87 | 0.74 | 1.02 |
| >= 2 Years and < 3 Years | 839 | 0.84 | 0.67 | 1.06 |
| >= 3 Years | 1,206 | 0.82 | 0.67 | 0.99 |
| Risperidone |  |  |  |  |
| Duration of Exposure |  |  |  |  |
| Never User | 709,280 | Reference | | |
| < 1 Month | 454 | 1.19 | 0.88 | 1.61 |
| >= 1 Month and < 3 Months | 2,326 | 1.32 | 1.16 | 1.52 |
| >= 3 Months and < 6 Months | 1,356 | 1.33 | 1.12 | 1.59 |
| >= 6 Months and < 9 Months | 831 | 1.50 | 1.21 | 1.88 |
| >= 9 Months and < 1 Year | 688 | 2.38 | 1.88 | 3.01 |
| >= 1 Year and < 2 Years | 1,383 | 2.04 | 1.72 | 2.42 |
| >= 2 Years and < 3 Years | 595 | 1.44 | 1.11 | 1.88 |
| >= 3 Years | 726 | 2.49 | 1.96 | 3.16 |
| Ziprasidone |  |  |  |  |
| Duration of Exposure |  |  |  |  |
| Never User | 716,612 | Reference | | |
| < 1 Month | 77 | 2.25 | 1.11 | 4.56 |
| >= 1 Month and < 3 Months | 317 | 2.31 | 1.64 | 3.25 |
| >= 3 Months and < 6 Months | 146 | 2.97 | 1.76 | 5.01 |
| >= 6 Months and < 9 Months | 85 | 2.39 | 1.25 | 4.58 |
| >= 9 Months and < 1 Year | 82 | 2.89 | 1.47 | 5.71 |
| >= 1 Year and < 2 Years | 143 | 3.35 | 2.00 | 5.61 |
| >= 2 Years and < 3 Years | 69 | 2.36 | 1.20 | 4.63 |
| >= 3 Years | 108 | 1.77 | 0.99 | 3.17 |

**Table S12: Population Attributable Risk for Use of Any Antipsychotic by Duration of Exposure.** Estimates were calculated using Levin’s formula with the exposure among the controls being used to estimate the expected exposure rate. Confidence intervals were calculated with 9,999 fractional weighted bootstraps to propagate uncertainty associated with the cross-duration-strata aggregation process. As with the marginal effects for the interaction models, weights were drawn from an Exp(1) distribution scaled to have a mean of 1 and the weights were constant within a stratum.

| Duration of Exposure | Strata Specific PAR | 95% CI |  |
| --- | --- | --- | --- |
|  |  | Lower Bound | Upper Bound |
| < 1 Month | -0.27 | -0.40 | -0.14 |
| 1-3 Months | 0.19 | 0.10 | 0.29 |
| 3-6 Months | 0.25 | 0.17 | 0.33 |
| 6-9 Months | 0.21 | 0.15 | 0.27 |
| 9-12 Months | 0.31 | 0.24 | 0.39 |
| 1-2 Years | 0.66 | 0.56 | 0.78 |
| 2-3 Years | 0.29 | 0.22 | 0.37 |
| 3+ Years | 0.66 | 0.55 | 0.77 |
| Any Exposure  (Sum of Above) | 2.26 | 1.98 | 2.57 |
